# Targeted T cell receptor gene editing provides predictable T cell product function for immunotherapy

**DOI:** 10.1101/2020.12.14.20248169

**Authors:** Thomas R. Müller, Sebastian Jarosch, Monika Hammel, Simon Grassmann, Manuel Effenberger, Immanuel Andrae, M. Zeeshan Chaudhry, Luka Cicin-Sain, Peter Steinberger, Michael Neuenhahn, Kilian Schober, Dirk H. Busch

## Abstract

Adoptive transfer of T cells expressing a transgenic T cell receptor (TCR) has the potential to revolutionize immunotherapy of infectious diseases and cancer. However, the generation of defined TCR-transgenic T cell medicinal products with predictable *in vivo* function still poses a major challenge and limits broader and more successful application of this ‘living drug’. Here, by studying 51 different TCRs, we show that conventional genetic engineering by viral transduction leads to variable TCR expression and T cell product functionality as a result of interference with the endogenous TCR, variable transgene copy numbers, and un-targeted transgene integration. In contrast, CRISPR/Cas9-mediated TCR gene editing enables defined, targeted TCR insertion with concomitant knock-out of the endogenous receptor. Thereby, T cell products display more homogenous and physiological TCR expression which results in increased functionality and – importantly – less variable *in vivo* T cell responses in comparison to conventionally generated T cells. Hence, targeted TCR gene editing increases the predictability of TCR-transgenic T cell product function, which represents a crucial aspect for clinical application in adoptive T cell immunotherapy.

**BRIEF SUMMARY:** Safer, more functional and predictable transgenic T cell products for immunotherapy can be generated via CRISPR/Cas9-mediated targeted TCR engineering

## Introduction

Adoptive T cell therapy (ACT) can specifically restore T cell-mediated immunity to fight infectious diseases or cancer (1, 2). The administration of autologous tumor-infiltrating lymphocytes (3) or transfer of pathogen-specific T cells that were *ex vivo* isolated from an HLA-matched donor (4) represent particularly successful approaches. However, while these approaches have proven their efficacy and safety, they are also largely restricted by the often limited availability and accessibility of antigen-specific, HLA-matched T cells from patient- or healthy donor-repertoires (5–7). This problem could be solved by genetically engineering TCR-transgenic T cells for clinical application since the desired TCR and the HLA-matched T cell can be taken from different sources. Furthermore, through combining optimal T cell phenotype (8–11), highest degree of HLA-matching (12), and TCR-intrinsic features such as antigen specificity or avidity (13, 14), TCR-engineered T cells provide unique therapeutic opportunities (15–17).

Yet, with regards to the TCR itself and its function after transgenic expression, many basic questions remain elusive. A major reason for this is the fact that transgenic re-expression of TCRs – a prerequisite for controlled investigation of TCR characteristics – is laborious. Very few studies have therefore investigated more than a handful of TCRs side-by-side (5, 18), and many basic questions about TCR biology have not yet been addressed. Moreover, even upon transgenic re-expression, the continued presence of the endogenous TCR and non-physiological TCR transgene expression after conventional editing of T cells (e.g. via viral transduction) hinder unbiased evaluation of TCR-intrinsic features.

Interactions between the endogenous and the introduced transgenic receptor can result in T cell products with inferior functionality (19, 20). The transgenic TCR competes with the endogenous TCR for a limited amount of CD3 molecules, which can decrease the surface expression of both (21). Furthermore, because of the TCR’s heterodimeric structure, chains of the transgenic and the endogenous receptor can form mispaired TCR variants, which again decreases surface expression of the desired transgenic TCR and also poses a safety hazard since mispaired TCRs can exert off-target toxicity (22–25). In the past, these problems in TCR engineering were tackled through many different approaches. Increased transgenic TCR surface expression and decreased levels of mispairing were achieved via murinization (26, 27) and an additional disulfide bond (28, 29) in the TCR constant regions, RNA-mediated silencing of the endogenous TCR (20), co-delivery of accessory or co-stimulatory molecules (30), introduction of single-chain TCRs (31), TCR constant region domain swapping (32) as well as framework engineering (18). However, complete elimination of mispairing and undisturbed expression of the transgenic TCR was only achieved via the full knock-out (KO) of the endogenous TCR (23, 33, 34).

In addition to the absence of the endogenous TCR, the quantity and site of transgenic TCR integration events represent crucial aspects for T cell product functionality and safety. Upon conventional transduction, advanced analysis of single-cell vector copy numbers (VCN) revealed up to 44 transgene integrations in a primary T cell (35). Both the US Food & Drug Administration (FDA) as well as the European Medicines Agency (EMA) highlight that the risk of gene-modified cell therapies via insertional oncogenesis (36) should be reduced through the limitation of VCN (37). Accordingly, clinical studies that reported VCN for antigen-specific receptor transgenic T cells documented an average VCN between 1 and 2 (38–40). However, even with low VCN, viral transduction results in close-to-random transgene integration (41) and requires constitutively active extrinsic gene promoters for transgene expression. While a low VCN may be desired for safety reasons, it may also limit TCR transcription levels and protein surface expression, which could ultimately compromise the functionality of conventionally engineered T cell products (20, 42, 43).

Novel advancements in the field of genetic engineering enable targeted TCR gene editing via KO of the endogenous TCR and simultaneous knock-in of a transgenic receptor into the respective TCR locus (44, 45). So-called orthotopic TCR replacement (OTR) results in the removal of endogenous TCR chains as well as targeted integration and more physiological regulation of the transgenic TCR (33).

In this report, we systematically compare OTR technology to conventional TCR editing in order to investigate how these methods affect the magnitude, variability, and interrelatedness of transgenic TCR surface expression and functionality. Furthermore, we assess the impact of interference with the endogenous TCR, low and high VCN, and defined versus random integration patterns on these parameters. Since TCR-intrinsic characteristics such as epitope-ligand specificity and avidity can bias general conclusions, we built up and used a library of 51 unique antigen-specific TCRs for this study. We noticed a large, yet TCR-intrinsic impact of competition and mispairing between endogenous and transgenic receptors. Interestingly, even after complete KO of the endogenous TCR, we observed robust functional differences between OTR and conventional TCR editing. We hypothesized that these differences originate from different and variable TCR transcription levels caused by random transgene integration and uncontrolled VCN after conventional editing. Indeed, in conventionally edited T cell products, we observed uncontrolled integration of TCR transgenes and substantial variability in TCR RNA expression on the single-cell level, which directly translated into heterogeneous TCR surface expression and variable functionality. In contrast to that, OTR resulted in homogenous and near-physiological expression of the transgenic TCR. Compared to conventional editing with a clinically relevant low VCN, the level of TCR surface expression through OTR was consistently higher, resulting in enhanced T cell functionality. Finally, the increased homogeneity of transgenic TCR expression after OTR led to more predictable *in vivo* T cell responses. These findings demonstrate that the method of TCR engineering can significantly shape T cell product functionality and recommend the usage of advanced, targeted gene editing tools for the production of clinically applied T cells. Targeted TCR gene editing via OTR offers not only a high genetic safety profile but also produces defined T cell products with increased and more predictable functionality compared to conventional editing.

## Results

### Variability of TCR engineered T cells through competition and mispairing

The study of TCRs and their engineering requires testing of more than one single receptor since individual TCR-intrinsic characteristics might bias general conclusions. Therefore, we developed a platform for reliable TCR sequence identification (Supplementary Fig. 1) and generated a library of Cytomegalovirus (CMV)-specific TCRs for in-depth functional characterization (Supplementary Table 1). To first validate the specificity and functionality of these TCR sequences in their native state, we generated fully human TCR constructs (i.e. not including any modifications such as murine TCR constant regions (26, 27)) for transgenic re-expression in primary human T cells via conventional retroviral transduction. Flow cytometry analysis of TCR-transgenic T cells revealed relatively poor TCR surface expression as indicated by low pMHC-multimer staining (Fig. 1a, 11.3% for TCR 15-11 and 2.54% for TCR 61-4). However, transgenic TCR surface expression was strongly increased, when an additional KO of endogenous TCR α- and β-chains (2xKO, KO-efficiency >93%) was performed (36.1% transgenic TCR expression for TCR 15-11 and 14.9% for TCR 61-4). We confirmed this observation with nine different A1/pp50_245-253_-specific TCRs, as indicated by an increased percentage of pMHC-multimer^+^ cells and pMHC multimer mean fluorescence intensity (MFI) after 2xKO (Fig. 1b). Interestingly, individual TCRs benefitted differentially from the additional 2xKO (as indicated by fold changes of the percentage of pMHC-multimer^+^ cells between ‘no KO’ and ‘2xKO’ ranging from 2.5 to 16.7).

**Fig 1.**
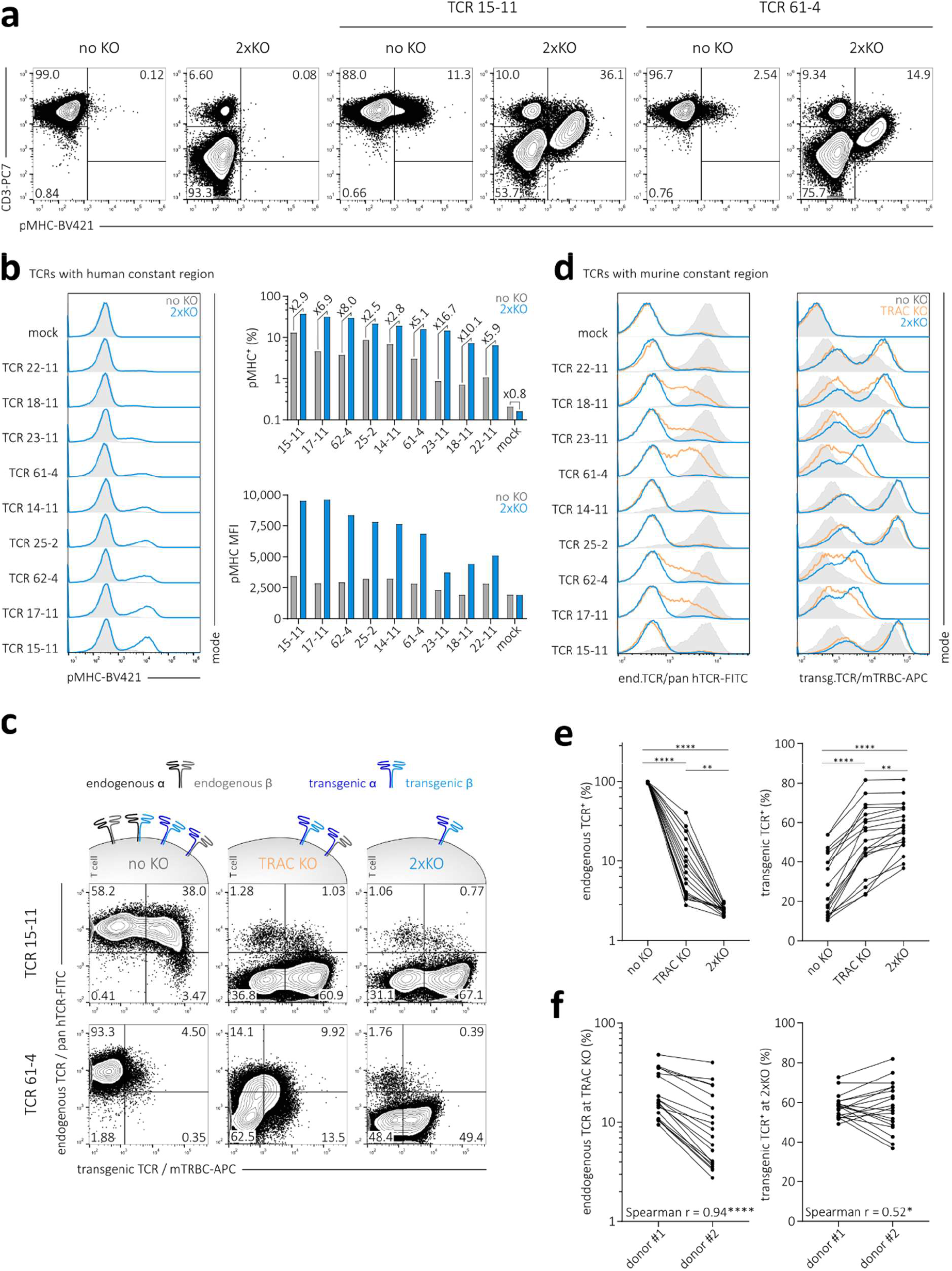
Variability of TCR engineered T cells through competition and mispairing. **(a)** Retroviral transduction of two human TCRs specific for A1/pp50 into human PBMCs with (2xKO, KO of TRAC and TRBC) or without (no KO) additional KO of the endogenous TCR. Numbers represent percentages of living CD8^+^ cells. **(b)** A1/pp50 pMHC-multimer staining (left), quantification of pMHC-multimer^+^ cells and fold change between editing methods for each TCR (top, right) as well as MFI of pMHC-multimer^+^ cells (bottom, right) after retroviral transduction of nine A1/pp50-specific fully human TCRs into PBMCs with (2xKO, blue) or without (no KO, grey) additional KO of the endogenous TCR. **(c)** Illustration (top) of possible interactions between endogenous and transgenic TCR in T cells without KO of the endogenous TCR (no KO, grey), KO of the TCR α-chain only (TRAC KO, orange) and KO of both TCR α- and β-chains (2xKO, blue). Flow cytometric analysis after co-staining (bottom) for endogenous and transgenic TCR for two representative A1/pp50-specific TCRs that contain a murine constant region (26). **(d)** Expression of endogenous TCR (left) and transgenic TCR (right) after retroviral transduction of nine A1/pp50-specific TCRs as in (c). **(e)** Percentage of CD8^+^ T cells expressing the endogenous TCR (left) and the transgenic TCR (right) for 19 different A1/pp50-specific TCRs (each represented by one dot per editing group). Statistical testing by one-way ANOVA (*** p<0.001) followed by Tukey’s multiple comparisons test, **** p<0.0001, ** p<0.01 **(f)** Percentage of CD8^+^ T cells expressing the endogenous TCR after retroviral TCR transduction and additional TRAC KO (left) and percentage of CD8^+^ T cells expressing the transgenic TCR after additional 2xKO (right) in two different donors. In both graphs, each dot represents one of 19 individual A1/pp50-specific TCRs per donor. Statistical testing by two-tailed Spearman correlation, **** p<0.0001, * p<0.05. Data are representative of two independent experiments.

We speculated that surface expression of transgenic TCRs without additional KO of endogenous chains might be decreased through competition for CD3 molecules with the fully paired endogenous TCR and through mispairing with individual chains of the endogenous TCR as reported by previous studies (21–24, 33, 46). Differences between transgenic TCRs may then be explained by differential competitiveness for CD3 molecules and/or dissimilar mispairing promiscuity. To investigate this, we generated TCR constructs with murine TCR constant regions (26) of 19 A1/pp50-specific TCRs (including the nine TCRs shown in Fig. 1b). Murinization of the transgenic TCR enables differentiation between the transgenic TCR and the endogenous TCR upon flow cytometric analysis via staining with an anti-murine TCR β-chain antibody (transgenic TCR / mTRBC) and a universal anti-human TCR antibody (endogenous TCR / pan hTCR) respectively (Fig. 1c). TCR-transgenic T cells without further editing (Fig. 1c, no KO) potentially express four different TCR variants: the full endogenous TCR, two mispaired variants, and the full transgenic TCR. In this setting, co-staining with mTRBC and pan hTCR visualizes T cells expressing the endogenous TCR only (mTRBC^-^ pan hTCR^+^, representing TCR un-edited T cells), T cells expressing both endogenous and transgenic receptor chains (mTRBC^+^ pan hTCR^+^) and T cells only expressing the transgenic TCR on the cell surface (mTRBC^+^ pan hTCR^-^). The double-positive mTRBC^+^ pan hTCR^+^ population shows a broad spectrum of both TCRs’ expression levels, indicating competition for CD3 and also variability between single T cells that might originate from different endogenous TCRs and their intrinsic competitiveness and mispairing promiscuity (46). Importantly, co-staining of cells with mTRBC and pan hTCR does not prove the existence of mispaired TCR variants in this setting. Only upon additional KO of individual TCR chains (Fig. 1c, TRAC KO), mTRBC and pan hTCR double-positive populations must originate from mispaired TCRs expressed on the cell surface. TCR 15-11 hardly shows such a double-positive population after TRAC KO, in sharp contrast to TCR 61-4, suggesting that mispairing promiscuity of individual transgenic TCRs is highly variable. As expected, mispairing can be completely eliminated by combinatorial KO of endogenous TCR α- and β-chains (Fig. 1c, 2xKO).

To further explore the scale and spectrum of TCR-intrinsic promiscuity for mispairing, we performed similar experiments with 19 A1/pp50-specific TCRs (see Fig. 1d for the same nine TCRs shown in Fig. 1b; see Supplementary Fig. 2a for remaining TCRs). TCRs with particularly suppressed transgenic TCR surface expression through mispairing in the TRAC KO setting (such as TCR 17-11, 62-4, 61-4) also showed a large gain in surface expression after 2xKO. Overall, we observed that surface expression of all 19 A1/pp50-specific TCRs significantly increased with additional editing of the endogenous TCR chains, but to a largely different extent for individual TCRs (Fig. 1e, right). This can be explained by the large spectrum of individual TCRs’ mispairing promiscuity as determined in the TRAC KO setting (Fig. 1e, left). Furthermore, TCR mispairing and gain in TCR surface expression correlate more strongly when T cells differ only in regard to mispairing (as seen by the comparison between 2xKO and TRAC KO T cells) than when T cells differ in regard to both mispairing and competition for CD3 molecules (as seen by the comparison between 2xKO and no KO; Supplementary Fig. 2b). This indicates that competitiveness for CD3 between correctly paired transgenic TCRs and endogenous TCRs is an additional, independent TCR-intrinsic feature that affects TCR surface expression. Since the endogenous TCR can in turn also be assumed to differ in its competitiveness for CD3 and its promiscuity for mispairing (46), we further aimed to investigate the role of the endogenous TCR repertoire of different T cell donors on expression and mispairing of transgenic TCRs. Interestingly, mispairing of individual transgenic TCRs was very similar between donors (Fig. 1f, left and Supplementary Fig. 3a), whereas TCR surface expression was remarkably different despite the full KO of endogenous TCRs (Fig. 1f, right and Supplementary Fig. 3b). In summary, individual transgenic TCRs – through their intrinsic differential competitiveness and mispairing promiscuity – introduce large variability in conventionally engineered T cell products. KO of endogenous TCR chains eliminates biases introduced by transgenic-endogenous TCR interactions, but inter-donor T cell product variability was still substantial.

### Targeted editing results in homogenous TCR expression and thereby reduces inter-donor T cell product variability

Intrigued by these results, we investigated potential reasons for the observed inter-donor TCR expression variability. Since interaction with the endogenous TCR as a confounding factor was eliminated, we speculated that the method of transgene integration could additionally contribute to the variability of TCR expression. Conventional editing, such as through lenti-/retroviral transduction or transposon systems, is a rather uncontrolled process during which one or multiple transgene copies integrate at least semi-randomly into the genome (47, 48). In contrast, OTR enables tightly controlled replacement of the endogenous TCR with transgenic TCR expression under the endogenous TCR promoter (33, 45). Therefore, we performed TCR insertion either via OTR or via retroviral transduction with PBMCs derived from three different donors (from here on always with simultaneous 2xKO of the endogenous αβTCR). For transduction, two different levels of virus MOI were used to achieve either integration of only one TCR copy according to Poisson-statistics (42) (Tx MOI^lo^) or a high copy number (Tx MOI^hi^). We observed that OTR generates lower percentages of transgenic TCR^+^ cells compared to retroviral transduction and that virus MOI correlates with editing efficiency (Fig. 2a,b). Notably, editing of five A2/pp65_495-503_-specific TCRs was least variable after Tx MOI^hi^ and most variable after Tx MOI^lo^ (Fig. 2b, see also Supplementary Fig. 4 for consistent results with TCRs with fully human constant regions). Despite the lower editing efficiency of OTR, inter-donor variability was significantly decreased in comparison to Tx MOI^lo^.

**Fig 2.**
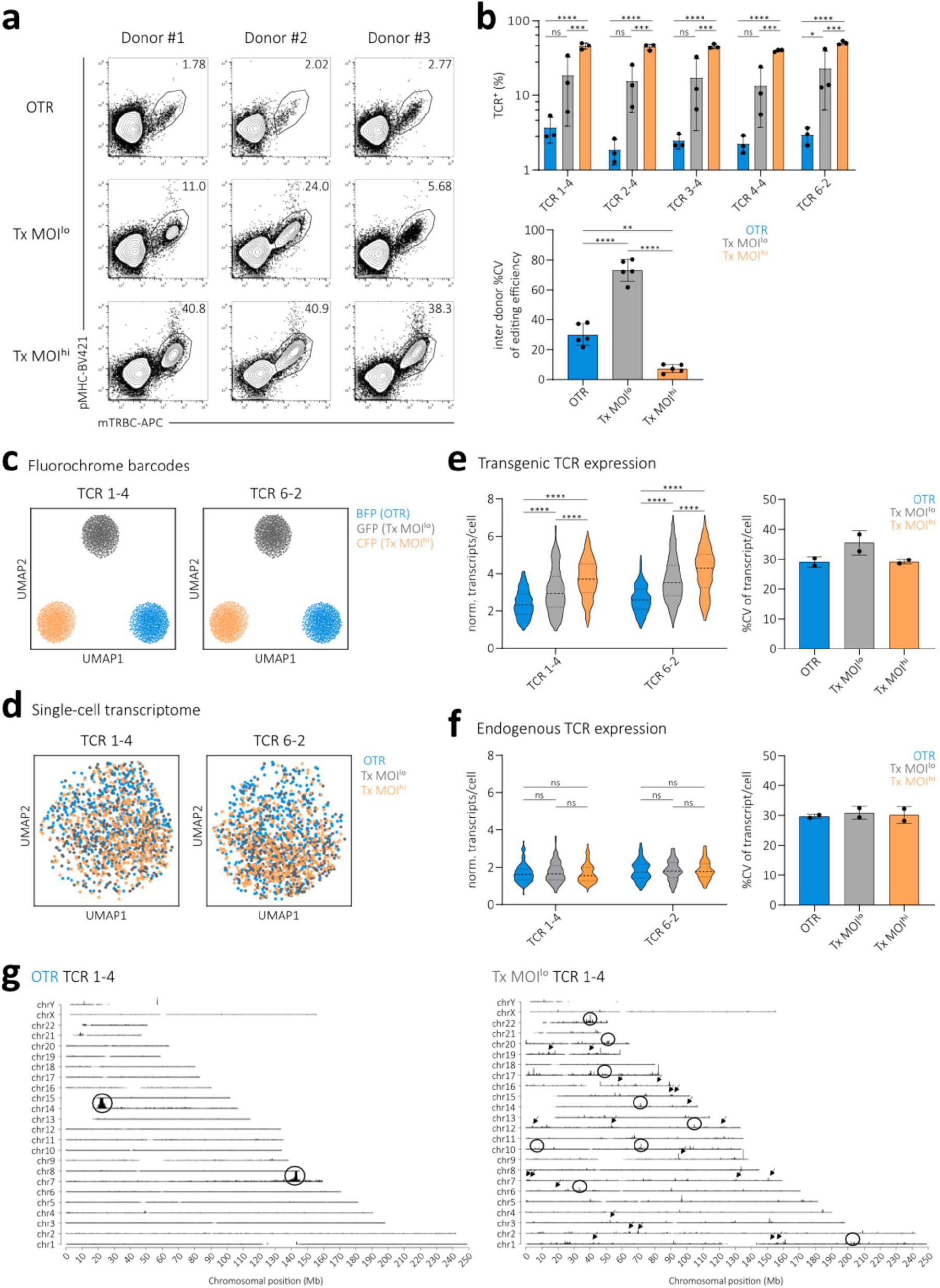
Targeted editing results in homogenous TCR expression and thereby reduces inter-donor T cell product variability. **(a)** Editing efficiency for one representative TCR specific for A2/pp65 in PBMCs of three different donors. Editing was performed either via OTR, retroviral transduction with a low virus MOI or with a high virus MOI as well as in all cases additional elimination of the endogenous TCR. Numbers denote percentages of CD8^+^. **(b)** Quantification of results from (a) for five A2/pp65-specific TCRs. Each dot represents the editing efficiency of an indicated TCR in one of three donors (top). Quantification of inter-donor variability for three different editing methods. Each dot represents inter-donor variability of one of five TCRs. Depicted is mean ± s.d.. Statistical testing by two-way ANOVA (**** p<0.001 for editing method, ns for TCR) followed by Tukey’s multiple comparisons test, **** p<0.0001, *** p<0.001, ** p<0.01, * p<0.05. Data are representative of two independent experiments. **(c)** Usage of fluorochrome barcodes for demultiplexing of samples. Visualization via uniform manifold approximation and projection (UMAP), n = 482 cells per barcode. **(d)** UMAP on whole transcriptome data compared by editing method for two A2/pp65-specific TCRs, n = 450 cells per editing method **(e**,**f)** Quantification of transgenic (e) and endogenous (f) TCR expression (left). Violin plots indicate frequency distribution, median and quartiles are shown with dashed lines. Statistical testing by two-way ANOVA ((e): **** p<0.001 for editing method, **** p<0.001 for TCR, n = 256 cells per editing method; (f): ** p<0.01 for editing method, ns for TCR, n = 108 cells per editing method) followed by Tukey’s multiple comparisons test, **** p<0.0001. Quantification of cell-to-cell variability (right). Depicted is mean ± s.d.. **(g)** TLA coverage across the human genome for integration of TCR 1-4. Circles indicate more abundant integration sites. Arrows indicate examples of less abundant integration sites. Similar results were obtained with an independent primer set.

We speculated that these observations on the protein level originate from transcriptional differences that can be related to the respective editing methods. Therefore, we performed single-cell RNA sequencing (scRNA seq) of TCR-edited T cells to measure transgenic as well as endogenous TCR expression in relation to the whole transcriptome. For this, we again performed OTR, Tx MOI^lo^, and Tx MOI^hi^ with two different TCRs. Additionally, we co-transduced three different fluorochromes (BFP, GFP, CFP), which served as barcodes for one of three different editing methods. This allowed us to multiplex all three editing samples of one TCR for sequencing in order to minimize biases through batch effects. Fluorochrome barcodes were successfully retrieved from sequencing data for both TCR pools and could be used for demultiplexing (Fig. 2c). In terms of the global transcriptome, we could not observe any substantial differences between editing methods (Fig. 2d) or between TCRs (Supplementary Fig. 5a). However, editing methods resulted in significantly different transgenic TCR expression (Fig. 2e) but consistent endogenous TCR expression (Fig. 2f). Of note, CRISPR/Cas9-mediated KO of endogenous TCR genes does not stop gene expression (so that they are still detectable via scRNA seq), but merely results in the production of dysfunctional and/or truncated proteins (33). In comparison to conventional editing, OTR resulted in a more homogenous TCR expression at a level that closely paralleled endogenous TCR expression. Within the two conventionally edited samples, higher virus MOI resulted in a higher transgenic TCR expression level as observed on the protein level. Moreover and analogous to the protein data shown in Fig. 2a and b, we observed increased transgenic TCR RNA expression variability after transduction with a low virus MOI in comparison to both OTR and Tx MOI^hi^ (Fig. 2e; Supplementary Fig. 5b), indicating defined transgene integration through OTR and synchronizing effects after transduction with a high virus MOI. House-keeping genes such as GAPDH, VIM, and MALAT1 were expressed uniformly irrespective of editing methods (Supplementary Fig. 5c-e).

The frequency distribution (as visualized by changing widths of violin plots in Fig. 2f) of transgenic TCR expression after Tx MOI^lo^ indicates that a fraction of cells might have received more than one transgene copy, which would be in line with previously reported single-cell VCN distributions (35). Another potential source of transgene expression variability is the genomic integration site itself. For example, different loci have different chromatin accessibility which affects gene transcription (49). To investigate integration sites, we performed targeted locus amplification (TLA) (50) with the same TCR re-directed T cell products shown in Fig. 2c-f. For OTR samples, we detected two integration sites (Fig. 2g left, Supplementary Fig. 6 left). Primarily, the full TCR transgene was inserted exactly at the intended target site in the first exon of the TCR α constant chain (TRAC) through homology-directed repair. In addition, homology-independent partial transgene integration occurred at the intended double-strand break in the TCR β constant region (TRBC1/2), as observed before (33). In sharp contrast to that, conventional editing resulted in a large number of integration sites dispersed over the whole genome. We detected 37 and 465 integration sites for TCR 1-4 Tx MOI^lo^ (Fig. 2g, right) and TCR 6-2 Tx MOI^lo^ (Supplementary Fig. 6, right) respectively. TLA indicated a high heterogeneity of the investigated T cell products so that many integration sites were below the detection limit. Hence, the detected number of integration sites most certainly represent underestimates. Bioinformatic analysis revealed that 27 (TCR 1-4 Tx MOI^lo^) and 196 gene regions (TCR 6-2 Tx MOI^lo^) were hit by transgene insertion (mostly intronic). Three (TCR 1-4 Tx MOI^lo^) and 35 (TCR 6-2 Tx MOI^lo^) integrations occurred in cancer genes. Interestingly, both Tx MOI^lo^ samples showed integrations into introns of the ‘cancer genes’ CHST11 and RUNX1. In summary, conventional editing introduces variability in TCR transgene and surface protein expression through random integration of an undefined transgene copy number per cell. This particularly affects clinically used low-virus dose transduced T cells since the genomic locus of a single integration site or the presence of a second transgene copy can have major consequences on TCR expression. In sharp contrast to that, defined TCR transgene integration via OTR results in homogenous and near-physiological TCR expression, which ultimately minimizes donor-dependent editing variability.

### Undefined VCN results in variable T cell product functionality

Having observed large editing variability after conventional editing with a low VCN, we speculated that this could also impact the functionality of TCR re-directed T cell products (20, 43). To test this hypothesis, we titrated virus MOIs for transduction of two A2/pp65-specific TCRs into endogenous TCR-KO T cells in order to simulate variable transgene integration numbers (Fig. 3a left panel and Supplementary Fig. 7a). As expected, editing efficiency and TCR surface expression positively correlated with virus MOI (Fig. 3a, left, and Supplementary Fig. 7a,b). We further observed that editing efficiency positively correlated with transgenic TCR surface expression (Fig. 3a, right). We then sorted by flow cytometry equal numbers of TCR-transgenic Tx MOI^lo^ (0.01x virus) and Tx MOI^hi^ (1.0x virus) CD8^+^ T cells and tested their functionality after a short time of *in vitro* culture. We could observe that increased transgenic TCR surface expression of Tx MOI^hi^ T cells resulted in significantly higher cytotoxic capacity (Fig. 3b) and peptide sensitivity (Supplementary Fig. 7c). To further corroborate our findings on the relationship between VCN, editing efficiency, TCR surface expression, and functionality, we introduced 36 A2/pp65-specific TCRs each into a Jurkat triple parameter reporter cell line (J-TPR) that allows for high throughput functional screening of TCR signalling (51). We observed that cells with high TCR surface expression (TCR^hi^) also showed increased levels of NFκB reporter activity upon antigen-specific stimulation (Fig. 3c). Maximum NFkB reporter activity in response to antigen (E_max_) of 36 TCRs was significantly increased in TCR^hi^ cells compared to TCR^lo^ cells (Fig. 3d). Peptide sensitivity was also significantly increased in TCR^hi^ cells (Supplementary Fig. 8a,b). In summary, editing with undefined VCN results in variable editing efficiency, surface expression, and T cell product functionality.

**Fig 3.**
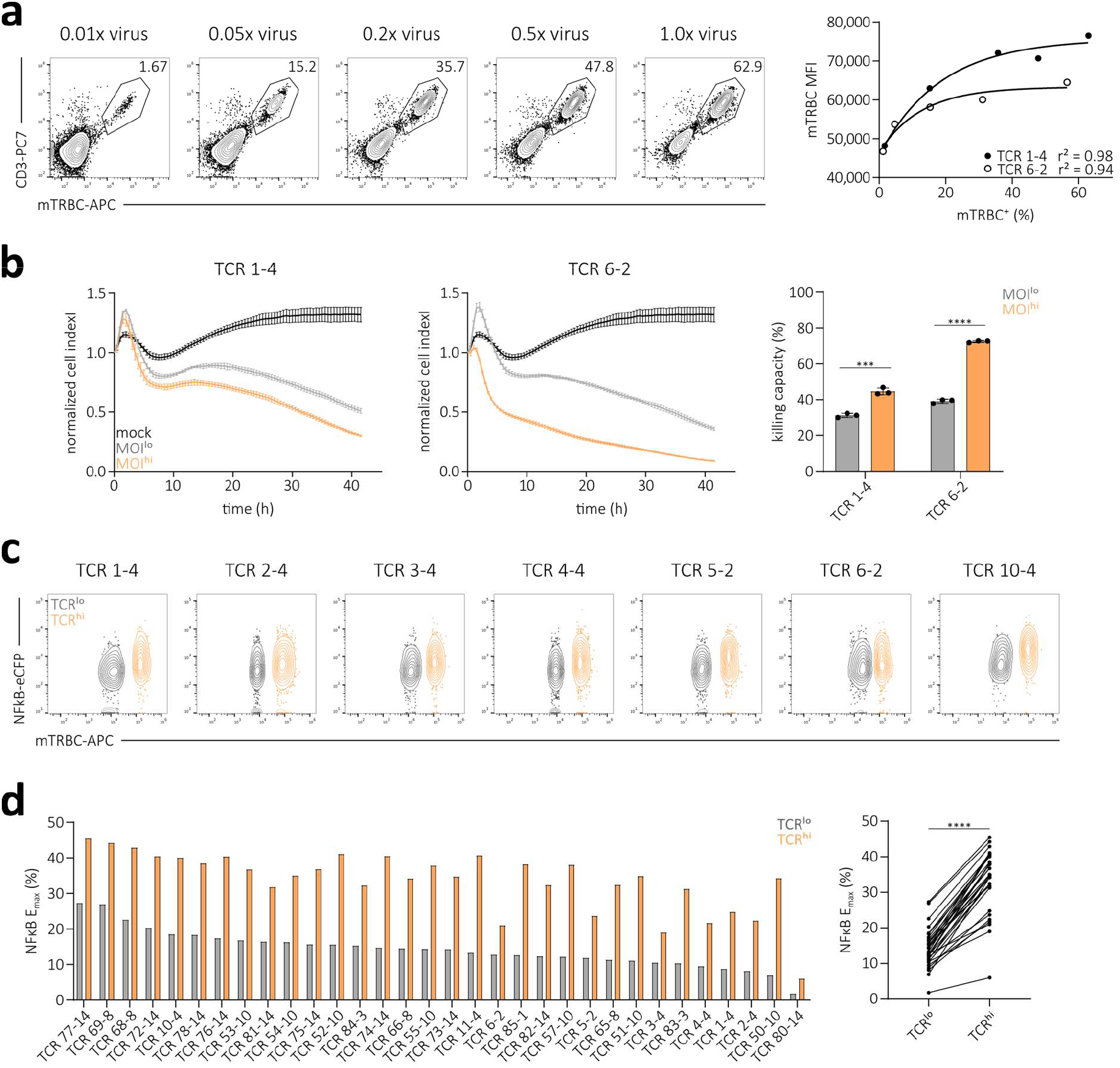
Undefined VCN results in variable T cell product functionality. **(a)** Transgenic TCR surface expression of the A2/pp65-specific TCR 6-2. Editing was performed in human PBMCs via retroviral transduction with a titration of virus dose and additional elimination of the endogenous TCR. Numbers indicate percentages of CD8^+^ (left). Correlation of editing efficiency to transgenic TCR surface expression (mTRBC MFI) (right). Each dot represents one of two indicated TCRs. Fitting by non-linear regression. **(b)** Killing of peptide-pulsed target cells over time by two A2/pp65-specific TCR-transgenic flow cytometry-sorted T cell products that were generated with a low (grey) or high (orange) virus MOI (0.01x virus and 1.0x virus, respectively; cell products depicted in (a) and Suppl. Fig. 7a). Lines illustrate the mean of three replicates ± s.d. (left). Quantification of killing capacity (as percentage of maximum killing, area under the curve normalized to mock control) for both TCRs and editing methods. Statistical testing by two-tailed unpaired Student’s t-test, **** p<0.0001, *** p<0.001 (right). **(c)** Flow cytometry measurement of TCR surface expression and NFκB reporter activity of seven representative A2/pp65-specific TCRs in the J-TPR cell line (51) after antigen-specific stimulation. Editing was performed via retroviral transduction and additional elimination of the endogenous TCR. **(d)** Maximum NFκB reporter activity in response to antigen (E_max_) of 32 A2/pp65-specific TCRs corresponding to data in (c). Bars represent mean of three replicates. Direct comparison between cells with low and high TCR surface (right). Statistical testing by two-tailed paired Student’s t-test, **** p<0.0001. Data are representative of two independent experiments.

### OTR generates defined T cell products with increased functionality

Antigen-specific receptor re-directed T cell products in past and present clinical trials have so far been generated via conventional editing with low virus MOIs^39–41^ in order to minimize risks originating from multiple (random) transgene integrations. We therefore set out to compare the functional capacity of T cells either generated via OTR or conventional editing with a low virus MOI. For this, we flow cytometry-sorted CD8^+^ transgenic TCR^+^ T cells and cultivated them in an antigen-free manner. While TCR RNA and protein expression levels were generally lower upon OTR a few days after editing (Fig. 2), we observed in several independent experiments and donor PBMCs that TCR surface expression levels were higher after OTR compared to transduction with low virus MOIs when cells were cultured for more than a week (Fig. 4a and Supplementary Fig. 9a). This possibly reflects a delayed accumulation of TCR surface protein after OTR, which is from then on maintained at a consistent level. Accordingly, after this culture period TCR surface expression was also more defined in OTR T cells, as indicated by narrower TCR staining peaks (Fig. 4a, left panel) and quantification of cell-to-cell surface expression variability (Supplementary Fig. 9b and 10). We then used these cells to study cytokine release upon antigen-specific stimulation and cytotoxicity. OTR T cells showed a more defined expression pattern of interferon-γ (IFNγ) and interleukin-2 (IL-2) (Fig. 4b, left panel) and a consistently enhanced peptide sensitivity in both donors and for all tested TCRs (Fig. 4b, right panel, Fig. 4c and Supplementary Fig. 9c). We also observed significantly higher cytotoxic capacities of OTR T cells (Fig. 4d). In summary, comparison of conventional TCR editing with a clinically relevant low copy number and OTR revealed increased TCR surface expression and functional capacity of OTR T cells.

**Fig 4.**
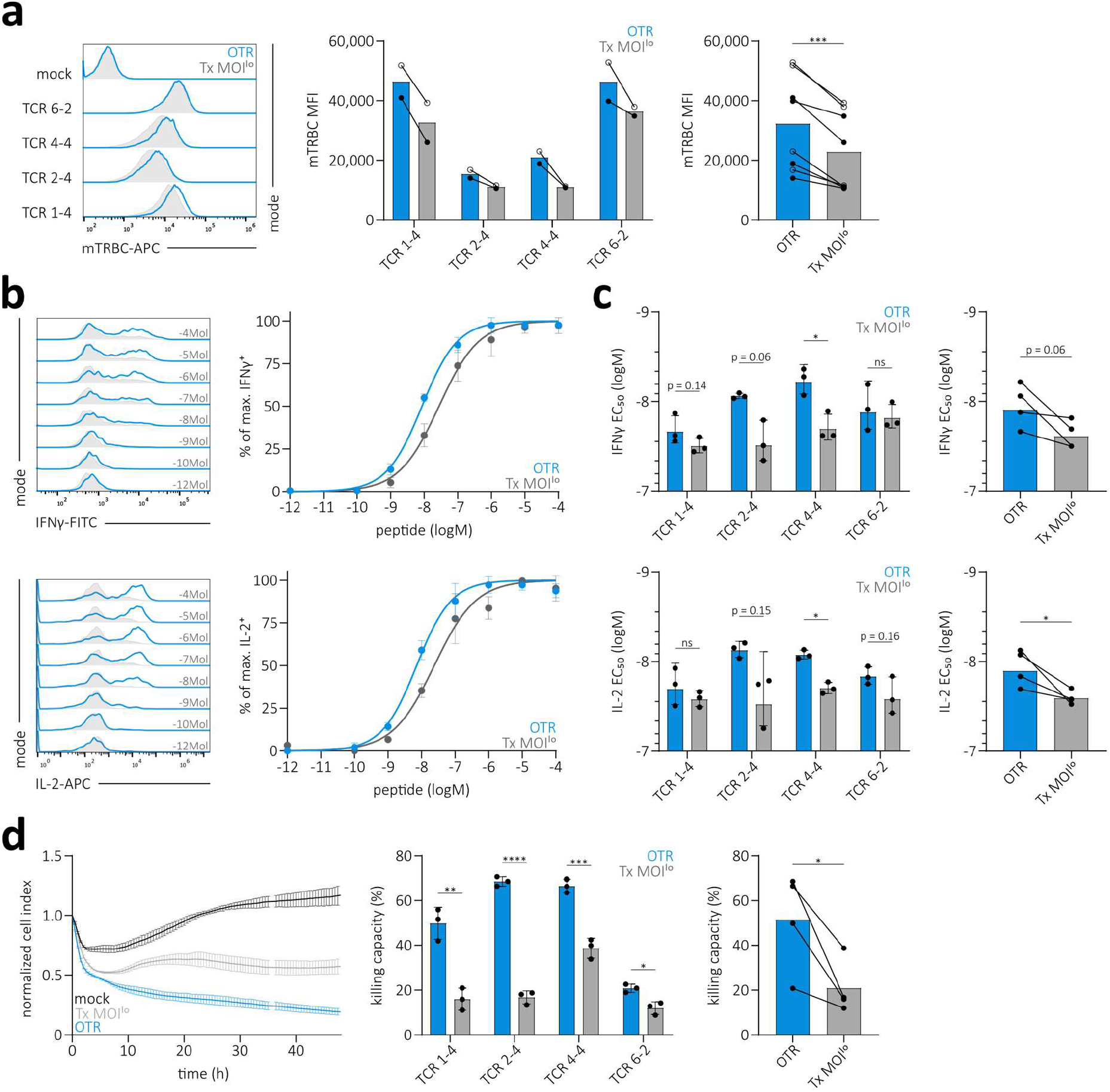
Uncontrolled TCR transgene integration results in variable surface expression and reduced T cell product functionality. **(a)** TCR surface expression of four A2/pp65-specific TCRs after flow cytometry-sorting and two weeks of *in vitro* culture. Editing was performed in human PBMCs via OTR (blue) or retroviral transduction with a low virus MOI (grey) and in both cases additional elimination of the endogenous TCR (left). Quantification of TCR surface expression levels for each TCR (middle) and pooled for comparison of editing groups (right). Each dot represents measurement of one TCR at one of two time points (day 10 after sort indicated by open circles, day 14 after sort by filled circles). Statistical testing by two-tailed paired Student’s t-test, *** p<0.001. **(b)** Functional cytokine response of T cell products shown in (a). Dose-dependent release of IFNγ (top left) and IL-2 (bottom left) after antigen-specific stimulation for one representative TCR. Corresponding EC_50_ curves (right). **(c)** Quantification of IFNγ (top left) and IL-2 (bottom left) EC_50_ values of four individual A2/pp65-specific TCRs. Depicted are replicates and mean ± s.d.. Statistical testing by two-tailed unpaired Student’s t-test. Direct comparison of IFNγ and IL-2 EC_50_ between editing groups (right). Here, each dot represents the mean EC_50_ of three replicates for one TCR. Statistical testing by two-tailed paired Student’s t-test, * p<0.05. **(d)** Killing capacity of T cell products shown in (a). Killing of peptide-pulsed target cells over time by four A2/pp65-specific TCR-transgenic T cell products. Lines illustrate the mean of three replicates ± s.d. (left). Quantification of killing capacity (as percentage of maximum killing, area under the curve normalized to mock control) for all four TCRs and editing methods (middle). Depicted are replicates and mean ± s.d.. Statistical testing by two-tailed unpaired Student’s t-test, **** p<0.0001, *** p<0.001, ** p<0.01, * p<0.05. Direct comparison between editing groups (right). Here, each dot represents the mean of three replicates for each TCR. Statistical testing by two-tailed paired Student’s t-test, * p<0.05.

### Targeted TCR gene editing results in more predictable *in vivo* T cell product function

We observed in multiple experiments with murinized TCRs (Fig. 2, Supplementary Fig. 9b and 10, four individual TCRs in three donors) as well as fully human TCRs (Supplementary Fig. 11, eight individual TCRs) that OTR generates highly defined TCR-transgenic T cell products in comparison to conventional editing. As we also collected evidence for a direct relationship between surface expression and *in vitro* functionality (see Fig. 3 and 4), we wondered whether OTR would result in more reliable *in vivo* functionality and thereby ultimately enable the generation of more predictable T cell products for immunotherapy.

Upon adoptive T cell transfer, not all transferred T cells are recovered and recruited into the immune response, as reflected by the recovery rate in single-cell transfer experiments (52). The transferred cells that ultimately form the immune response are therefore a subpopulation of the initially transferred T cell product. Accordingly, we hypothesized that the variable TCR expression between single cells of a transferred T cell product should also lead to functional variability between recruited cell populations in different hosts. To investigate this, we introduced four different fully human A2/pp65-specific TCRs into T cells either via OTR or conventional editing. In addition to TCR editing, we further introduced four different fluorochrome transgenes in order to color-barcode each TCR (53). This allowed us to perform polyclonal adoptive transfer experiments, thereby reducing technical mouse-to-mouse variability (54). We pooled all four TCR-transgenic T cell products that were either generated via OTR or conventional editing and transferred the cells (1 x 10^5^ cells per TCR) together with human IL-2 via intraperitoneal injection (i.p.) into irradiated NSG/HHD HLA-A*02 mice that were subsequently infected with a transgenic murine CMV (mCMV) strain that presents the human CMV (hCMV) HLA-A*02-restricted pp65 epitope (NLV) (Fig. 5a). This model allows for *in vivo* investigation of human TCR-transgenic T cells in a system with natural CMV tropism (55). As observed before (Supplementary Fig. 9b, 10, and 11), fluorescence intensity (FI) of individual cells after pMHC-multimer staining was significantly less variable in OTR T cells (Fig. 5b). Using data from Fig. 5b, we simulated the effect that a 10% *in vivo* recovery rate would have on the composition of resulting populations (Fig. 5c). Random draws of ten from 100 representative TCR-transgenic cells generated populations with either minor (OTR) or substantial differences in TCR expression (Tx), both in terms of FI variability as well as in terms of MFI variability (Fig. 5c, upper panel). FI variability thereby reflects TCR surface expression within a population, while MFI variability reflects the variability of mean TCR surface expression levels between different populations. We performed 100 of such random 10-cell draws and calculated the TCR expression variability in each of the 100 generated populations, which illustrates reduced variability after OTR (Fig. 5c, lower left panel). Calculated MFIs of all 100 individual populations were also less variable after OTR compared to conventional editing (Fig. 5c, lower right panel), which should reduce variability of inter-host T cell responses. In order to experimentally test the functional outcome of this simulation, we measured T cell responses of the four different TCR-transgenic T cell products that were transferred into individual mice. The additional fluorochrome color-barcode enabled us to differentiate between TCRs so that we could compare individual TCR population sizes between different mice (Fig. 5d, for gating strategy see Supplementary Fig. 12a). We observed that inter-host functional variability of individual TCRs was smaller when T cells were produced via OTR compared to conventional editing (Fig. 5e and Supplementary Fig. 12b). To corroborate this finding, we further performed a second experiment with four – now murinized - A2/pp65-specific TCRs and again observed significantly reduced functional inter-host variability of OTR T cells (Fig. 5f and Supplementary Fig. 12c). In summary, TCR-transgenic T cell products generated through targeted TCR gene editing showed reduced cell-to-cell TCR surface expression variability and, most importantly, through this a more predictable *in vivo* functional response compared to conventional editing.

**Fig 5.**
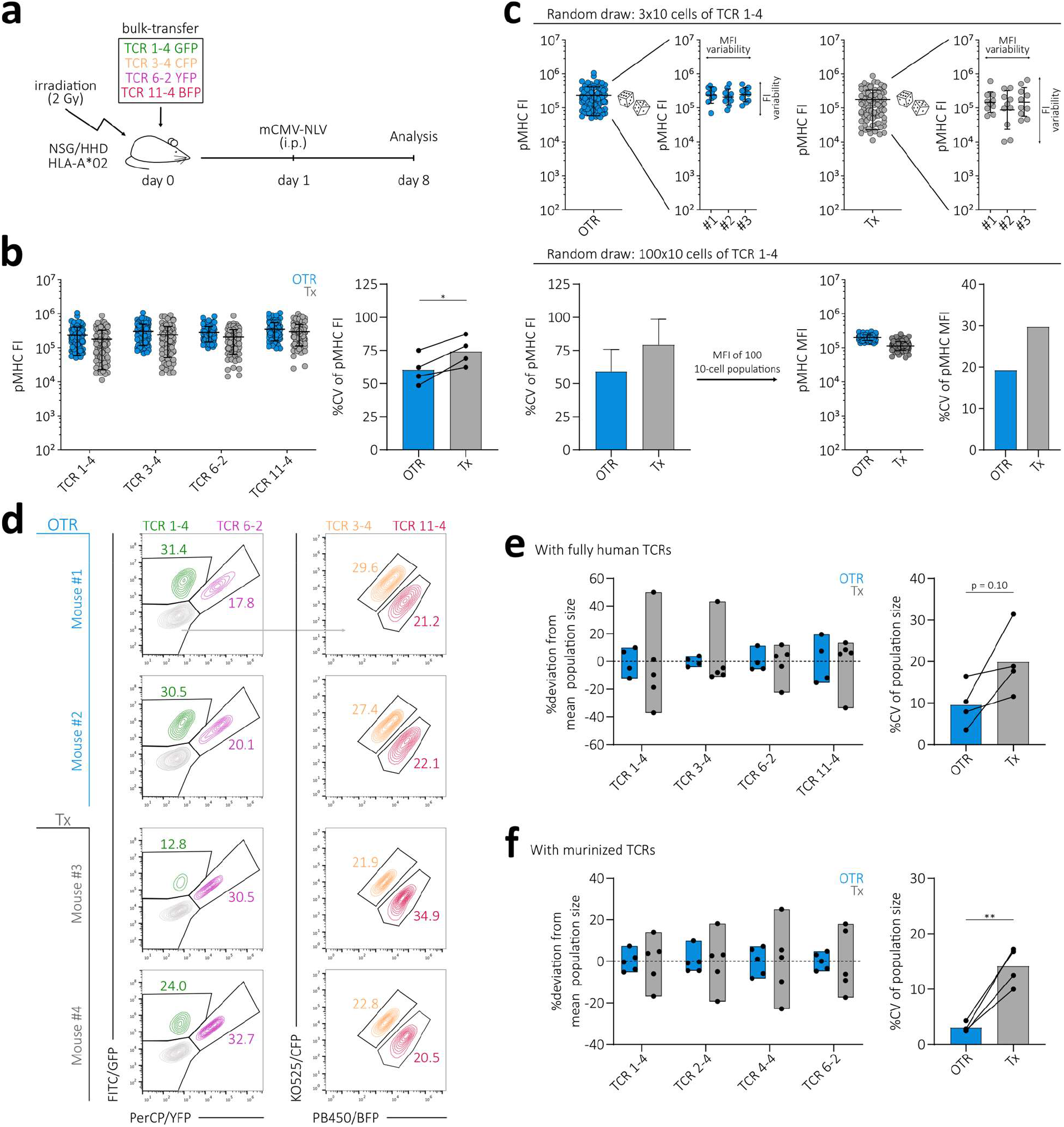
Targeted TCR gene editing results in more predictable *in vivo* T cell product function. **(a)** Schematic illustration of *in vivo* experiment. **(b)** pMHC-multimer fluorescence intensity (FI) of four fully human A2/pp65-specific TCRs. Editing was performed either via OTR (blue) or retroviral transduction (grey) and additional elimination of the endogenous TCR (accounts for both methods). Each dot represents one single cell. For each TCR and editing method, 100 randomly drawn cells derived from a larger population are displayed (left). Quantification of the corresponding cell-to-cell pMHC-multimer FI variability (right). Statistical testing by two-tailed paired Student’s t-test, * p<0.05. **(c)** Simulation of surface expression after a 10% *in vivo* recovery rate by random draws of 10 cells from the 100-cell population of TCR 1-4 shown in (b) (top panel). Analysis of cell-to-cell pMHC FI variability in 100 randomly drawn 10-cell populations (bottom left) and their corresponding MFIs as well as variability between 100 MFIs (bottom right). **(d)** Analysis of TCR-transgenic T cell responses at day 8 in the liver of sacrificed animals. Depicted are representative responses in two mice per editing group. Numbers denote percentages of human CD8^+^pMHC^+^ cells. **(e)** Quantification of TCR-transgenic T cell response variability between individual mice. For each TCR and editing method, the percentage deviation of total T cell recruitment in one mouse compared to the mean recruitment in all mice is depicted as one dot (left). Bars indicate minimal to maximal deviation from mean. Quantification of inter-host variability of T cell responses by editing group (right). Each dot represents one TCR. Statistical testing by two-tailed Student’s t-test; n = 4-5 mice per group. **(f)** Repetition of the experiment shown in (e) with four murinized A2/pp65-specific TCRs in PBMCs of different donor origin than shown in (e). Statistical testing by two-tailed Student’s t-test, ** p<0.01; n = 5 mice per group.

## Discussion

First reports on CRISPR/Cas9-mediated replacement of the endogenous TCR with either a chimeric antigen receptor (CAR) (44) or a TCR (45) were perceived as important proof-of-concept studies in the fields of T cell engineering and ACT. Targeted transgene integration not only promised to offer an increased safety profile but is also highly appealing as OTR T cell products should very closely resemble physiological, unedited T cells. Indeed, placing an antigen-specific receptor under endogenous TCR transcriptional control facilitates near-physiological TCR regulation (33). Despite these advancements, many questions on how OTR T cell products differ from conventionally edited T cells have remained unresolved. Yet, this comparison is of fundamental importance for basic research on TCR biology as well as for the field of ACT given that conventional editing via viral transduction is currently the most widely used method for the production of clinically applied T cell products. In this study, we re-expressed 51 antigen-specific TCRs via OTR and conventional editing and investigated the consequences of differential genetic TCR integration profiles on the magnitude, variability, and interrelatedness of transgenic TCR surface expression and functionality.

We first validated a large, yet TCR-intrinsic impact on competition and mispairing between endogenous and transgenic receptors on TCR expression. The concept of ‘weak’ and ‘strong’ recombinant TCR expression is well established (46, 56). A recent study showed that a few amino acids in the TCR variable regions have a great impact on TCR surface expression, potentially due to increased protein folding efficiency and/or α-β-chain assembly (18). However, addressing whether strong TCRs are more capable of outcompeting endogenous TCRs, or whether they are less affected by mispairing of individual TCR chains, is only possible through genetic KO studies. Our results indicate that TCR competitiveness and mispairing promiscuity are independent and TCR-intrinsic features, which both affect transgenic TCR surface expression. We could further reveal that mispairing promiscuity is not a digital but an analogous event. In the future, it will be interesting to investigate how these two factors are separately coded within the TCR sequence. Overall, our data underline the importance of endogenous TCR-KO in order to generate safer and more standardized TCR-redirected T cell products.

Intriguingly, even upon full KO of the endogenous TCR, we still observed substantial inter-donor variability after conventional editing. We speculated that this variability could originate from random integration of an undefined number of transgene copies. In fact, it is well known that VCN affects editing efficiency and transgene expression (20, 42). Clinically applied, conventionally edited TCR re-directed T cell products showed sometimes promising, yet overall rather variable therapeutic results (15–17, 40). This might be – at least to some extent – attributable to uncontrolled TCR transgene integration.

Here, we measured TCR transgene integration site, transcription, surface expression, and T cell product functionality after conventional editing with a low or high virus MOI, as well as after OTR. Thereby, we could directly relate defined transgene integration via OTR to a more homogenous, physiological TCR transcription as well as to a less variable surface expression and functionality. In case of conventional editing, both variable VCN and random transgene integration independently increase the variability of TCR transcription, surface expression, and functionality – especially after transduction with a low virus MOI. On the one hand, this can be explained by VCN heterogeneity between single cells (35). On the other hand, differential accessibility of a transgene at a specific genomic locus should have a large impact on transgene transcription (49). In this regard, variability should be amplified when only a single transgene integration took place (as with low virus MOI), whereas, upon multiple integrations, averaging effects can synchronize TCR expression (as with high virus MOI). This is ultimately reflected in the higher inter-donor variability of TCR surface expression in MOI^lo^ edited cells. Most importantly, the level of TCR surface expression directly affects functionality, which implicates that conventional editing with a low virus MOI generates T cell products of variable function. Such variability can be decreased through transduction with a high virus load. However, the random integration pattern of conventional editing in combination with a high VCN in turn increases the risk of insertional oncogenesis as observed before in a clinical trial (57). Consequently, the use of high VCN for the generation of transgenic T cell products is discouraged by FDA and EMA (36).

Concomitantly, controlling for a defined VCN standardizes the functional comparison between editing methods. This aspect has not been taken into account by previous studies, which reported that OTR T cells exhibit similar functionality *in vitro* (33, 45, 58) and similar or enhanced functionality *in vivo* (45, 58). Here, we show that all four tested A2/pp65-specific TCRs showed increased TCR surface expression and functionality when introduced via OTR compared to transduction with a low virus MOI. In the future, long-term *in vivo* protection experiments are needed to test whether OTR T cells show prolonged maintenance and enhanced effector function, as proposed by more physiological TCR regulation (33) and OTR CAR-T cell data (44).

We observed by scRNA seq as well as flow cytometry that OTR decreases cell-to-cell TCR expression variability. In contrast, conventional editing results in variable TCR surface expression that directly affects functionality. Using a novel polyclonal transfer system to study different color-barcoded TCRs side-by-side *in vivo*, we could show that this observed heterogeneity within the T cell product introduces substantial T cell response variability. For clinical application, a highly defined and homogenous T cell product that provides predictable *in vivo* function is of utmost importance. Targeted TCR gene editing via OTR facilitates the production of such highly defined T cell products with an enhanced safety profile as well as increased and predictable functionality.

## Material & Methods

### T cells from PBMCs and cell culture

T cells were cultured in RPMI 1640 (Gibco) supplemented with 10 % FCS, 0.025% L-glutamine, 0.1% HEPES, 0.001% gentamycin and 0.002% streptomycin and 180 IU ml^-1^ IL-2 (‘RPMI’ hereafter) unless indicated otherwise. Sorted cells were cultured with 1×10^6^ ml^-1^ irradiated (30 Gy) allogeneic feeder PBMCs, 1 µg ml^-1^ PHA and 180 IU ml^-1^ IL-2.

Written informed consent was obtained from the donors, and usage of the blood samples was approved according to national law by the local Institutional Review Board (Ethikkommission der Medizinischen Fakultät der Technischen Universität München).

### TCR identification

CMV-seropositive donor PBMCs were stained with respective pMHC-multimer that was individually conjugated with two different fluorophores to achieve reliable double pMHC-multimer staining. Single cells positive for CD8, CD62L, CD45RO and pMHC-multimer were sorted in a 384 well plate and stimulated with 10µg ml^-1^ plate bound anti-CD3 and anti-CD28 each. RPMI medium was supplemented with 200 IU ml^-1^ IL-2 and 5 ng ml^-1^ IL-15. Single cell derived clones were harvested between day 7 and 14 after sort. TCRs were amplified via TCR-SCAN RACE PCR (59) and subsequently sequenced on the Illumina MiSeq platform.

### TCR DNA template design

DNA templates were designed *in silico* and synthesized by GeneArt (Life Technologies, Thermo Fisher Scientific) or Twist Bioscience. DNA constructs for retroviral transduction had the following structure: Human Kozac sequence (60) followed by TCR β (including as indicated either human TRBC or murine TRBC with additional cysteine bridge (26, 28, 29)), followed by P2A, followed by TCR α (including as indicated either human TRAC or murine TRAC with additional cysteine bridge (26, 28, 29)), cloned into pMP71 vectors (kindly provided by Wolfgang Uckert, Berlin). DNA constructs for CRISPR/Cas9-mediated homology-directed repair (HDR) had the following structure: 5’ homology arm (300-400 bp), P2A, TCR β (as above), T2A, TCR α (as above), bGHpA tail, 3’ homology arm (300-400 bp). The homology arm sequences of the TRBC locus were derived from TRBC1 and are highly homologous to TRBC2.

### CRISPR/Cas9-mediated KO and KI

Cas9 RNPs and HDR repair templates were generated as described before (33). Either column selected CD4^+^ and CD8^+^ T cells mixed in a 1:1 ratio or bulk PBMCs were activated for two days in RPMI with CD3/CD28 Expamer (Juno Therapeutics), 300 IU ml^-1^ IL-2, 5 ng ml^-1^ IL-7 and 5 ng ml^-1^ IL-15. Expamer stimulus was removed by incubation with 1 mM D-biotin. Cells were electroporated (pulse code EH100) with Cas9 ribonucleoprotein and DNA templates in Nucleofector Solution (20 µl per 1 x 10^6^ T cells; Lonza) with a 4D Nucleofector X unit (Lonza). After electroporation, cells were cultured in RPMI with 180 IU ml^-1^ IL-2 until a first FACS analysis on day five after editing.

### Retroviral transduction

For the production of retroviral particles, RD114 were transfected with pMP71 expression vector (containing a TCR or flourochrome) by calcium phosphate precipitation. Virus supernatant was coated on retronectin (TaKaRa) treated well plates. Either column selected CD4^+^ and CD8^+^ T cells mixed in a 1:1 ratio or bulk PBMCs were activated for two days in RPMI with CD3/CD28 Expamer (Juno Therapeutics), 300 IU ml^-1^ IL-2, 5 ng ml^-1^ IL-7 and 5 ng ml^-1^ IL-15. Expamer stimulus was removed by incubation with 1 mM D-biotin. Activated T cells were transduced via spinoculation on virus-coated plates. TCR and/or fluorochrome transduction occurred 15 min after CRISPR/Cas9-mediated TCR KO editing.

### pMHC multimer and antibody staining

pMHC monomers were generated as previously described (61). All pMHC-multimer reagents were generated by incubation of 4 µg biotinylated pMHC monomer with 1 µg streptavidin-BV421 in a total volume of 100 µl FACS buffer for staining of up to 1 x 10^7^ cells. The following antibodies were used: anti-human TCR α/β FITC (BioLegend), CD3 PC7 (BD Biosciences), CD8 PE (Invitrogen), anti-murine TCR β-chain APC (BioLegend), CD62L FITC (BD Biosciences) and CD45-RO PC7 (BioLegend). Live/dead discrimination was performed with propidium iodide (Invitrogen).

### Antigen-specific activation and intracellular cytokine staining

One the day before co-culture with T cells, K562 cells (retrovirally transduced to express the human MHC class-I molecule of interest) were irradiated (80 Gy) and loaded with peptide (10^−12^ M, 10^−10^ M, 10^−9^ M, 10^−8^ M, 10^−7^ M, 10^−6^ M, 10^−5^ M, 10^−4^ M) overnight at 37°C. T cells were co-cultured with peptide-loaded K562 cells and Golgi plug (BD Biosciences) in a 1:1 ratio for 4 h at 37°C. PMA (25 ng ml^-1^) and ionomycin (1µg ml^-1^) were used for positive control. pMHC-multimer and surface marker antibody staining for CD8 (PE, Invitrogen) and anti-murine TCR β-chain (APC/Fire750, BioLegend) was followed by permeabilization using Cytofix/Cytoperm (BD Biosciences), and intracellular staining of IFNγ (FITC, BD Pharmingen), TNFα (PC7, eBioscience) and IL-2 (APC, BD Biosciences). Live/dead discrimination was performed with ethidium-monoazide-bromide (Invitrogen).

### Generation and analysis of TCR-edited human T cell reporter lines

TCRs were introduced into the J-TPR cell line (51) via retroviral transduction. Antigen-specific stimulation was performed using irradiated (80 Gy) and peptide pulsed (10^−9^ M, 10^−8^ M, 10^−7^ M, 10^−6^ M, 10^−5^ M, 10^−4^ M) K562 cells (retrovirally transduced to express the human MHC class-I molecule of interest). Effector and target cells were co-cultured in a 1:5 ratio for 18 h. Subsequently, NFκB-eCFP reporter activity of J-TPR cells was analysed on a flow cytometer.

### Cytotoxic T lymphocyte assay

HLA-A*0201 positive HepG2 cells were loaded with 10^−10^ M of A2/pp65495-503 (NLV) peptide. 4 x 10^4^ peptide pulsed HepG2 cells were plated per well in a 96 well E-Plate (ACEA Biosciences). The plate was subsequently placed into a xCELLigence RTCA MP Real Time Cell Analyzer (ACEA Biosciences) and HepG2 cell growth was monitored every 15 minutes. After 24 hours, 4 x 10^4^ rested T cells were added per well containing HepG2 target cells. Mock edited (TCR transgene negative) T cells derived of the same donor were used as negative control. Effector and target cells were co-incubated for 48 h and cell growth/death was monitored every 15 minutes.

### Flow cytometry

Acquisition of FACS samples was done on a Cytoflex (S) flow cytometer (Beckman Coulter). Flow sorting was conducted on a FACSAria III (BD Bioscience) or MoFlo Astrios EG (Beckman Coulter).

### scRNA sequencing

TCRs were introduced either via CRISPR/Cas9-mediated KI or via retroviral transduction into endogenous TCR-KO primary T cells as described. T cells that underwent different TCR editing approaches (OTR, MOI^lo^, MOI^hi^) were barcoded via co-transduction with three different fluorochromes (BFP, CFP, GFP). Five days after editing, FACS sorted CD8^+^TCR^+^fluorochrome^+^ cells were used for determination of transgenic TCR expression and whole transcriptome analysis on the 10x Genomics platform. Protocol was performed according to manufacturer’s instruction. Only adaptation was the application of custom primer sets for specific amplification of transgenic TCR constructs.

### scRNA sequencing – Data processing

Combined fastq-files (transcriptome and transgenic TCR library) for each TCR were annotated against a custom reference containing all genes of the human genome (GRCh38), the fluorochromes (BFP, CFP, GFP) used for multiplexing and the respective TCR constructs using Cell Ranger (V3.0.2). Data analysis of the annotated count matrix was performed in SCANPY (62). Cells were filtered to contain at least 200 genes and genes being present in less than 3 cells were excluded. 20% mitochondrial gene expression was allowed. Counts were normalized to 10.000 counts per cell and expression was log transformed. The number of counts, percent of mitochondrial genes, S and G2M phase scores were regressed out. In order to demultiplex subsamples according to fluorochrome expression, cells were filtered to express only one of the three fluorochromes and resulting leiden-clusters (63) of the neighbourhood graph were annotated according to fluorochrome expression. Highly variable genes (HVGs) were identified with mean values between 0.0125 and 3 and a minimal dispersion of 0.5. Expression values exceeding a standard deviation of 10 were clipped. The neighbourhood graph was calculated for the 10 nearest neighbours and the first 7 components of the PCA for the HVGs. Fluorochromes and constructs have been excluded for the neighbourhood embedding. Violin plots show the normalized raw gene expression.

### Targeted locus amplification

TLA including sequencing was performed by Cergentis as previously described (50). Two primer sets were designed to target each transgene. Primer sets were used in individual TLA amplifications. PCR products were purified and library prepped using the Illumina Nextera flex protocol and sequenced on an Illumina sequencer. NGS reads were aligned to the transgene sequence and host genome (human hg19 reference sequence). Bioinformatic analysis of integration sites and detection of hit cancer genes via Enhort (unpublished, https://enhort.mni.thm.de/), Cosmic (64) and Network of Cancer Genes (65).

### mCMV mutagenesis

MCMV-ie2-ANLV was generated by fusing AANLVPMVATV peptide at the C-terminus of the ie2 protein. HCMV pp65_495-503_ epitope (NLVPMVATV) was preceded by two alanine residues that enhance peptide processing and presentation (66). The peptide was inserted at the ie2 C-terminus position 187,296 (NCBI accession: NC_004065) using en passant mutagenesis as described before (66, 67).

### *In vivo* assay

*In vivo* experiments were performed with HLA-A*0201 transgenic NOD/SCID/IL-2rg^-/-^ mice and a chimeric murine CMV engineered to express the human CMV A2/pp65_495-503_ (NLV) peptide (55). First, mice were irradiated (2 Gy) and subsequently 1 x 10^5^ CD8^+^TCR^+^ cells (per respective TCR) were injected intraperitoneally. About 24 h after infection, mice were infected with 5 x 10^3^ PFU of chimeric mCMV. Mice were sacrificed at day 9 after T cell transfer and their liver harvested. Intra-hepatic lymphocytes were subsequently stained with pMHC-multimer, anti-human CD45 PC7 (eBiosciences), anti-human CD8 PE (Invitrogen), anti-murine TCR β-chain APC/Fire750 (Biolegend) and propidium iodide (Invitrogen).

### Data analysis

All data were analysed with FlowJo v10 and GraphPad PRISM software.

## Data Availability

All data generated or analyzed during this study are included in this article and its supplementary information files. Additional raw data are available from the corresponding authors upon reasonable request.

## Acknowledgments

We thank members of the Busch and Buchholz laboratories for experimental help and critical discussion, particularly F. Mohr, J. Leube, M. Plambeck, A. Hochholzer, F. Graml, S. Dötsch, E. d’Ippolito and V. R. Buchholz. We also thank our flow cytometry unit, specifically L. Henkel, C. Angerpointner and M. Schiemann. We appreciate fruitful discussions with C. Stemberger, M. Poltorak and L. Germeroth (Juno Therapeutics). We are also grateful to O. Quitt, U. Protzer, K. Dennehy and W. Uckert for providing cell lines and constructs. This work was mainly supported by the German Center for Infection Research (DZIF).

## Author contributions

T.R.M., K.S. and D.H.B. conceived the study. T.R.M., K.S. and D.H.B. designed and analysed experiments. T.R.M., M.H. and K.S. developed the TCR library. M.N. provided a donor biobank for TCR isolation. M.E. generated pMHC-multimers. T.R.M performed TCR editing and *in vitro* experiments. P.S. developed and advised on experiments with J-TPR cell line. T.R.M and K.S. performed *in vivo* experiments. M.Z.C and L.C-S. generated mCMV for *in vivo* experiments. S.G. and I.A. performed and advised on FACS sorting. T.R.M., S.J. and M.H. performed single-cell transcriptomics. S.J. performed bioinformatics analysis. T.R.M., K.S. and D.H.B. wrote the manuscript. All authors read and reviewed the manuscript.

## Supplementary Figures

**Suppl. Fig 1.**
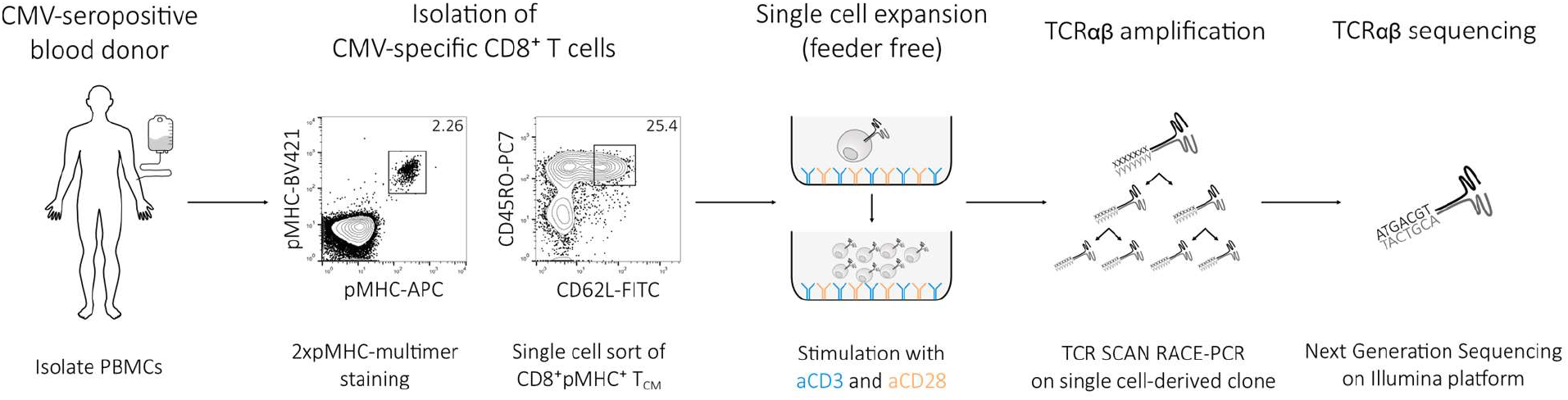
A platform for reliable TCRαβ sequence identification from virus-seropositive donors. A schematic illustration of the TCRαβ sequence identification workflow, from left to right: Isolation of PBMCs from CMV-seropositive blood donors. Staining of PBMCs with double pMHC-multimer (same peptide-MHC monomer, but backbones labelled with different fluorochromes) and T cell memory markers (pre-gated on living CD3^+^CD8^+^CD19^-^). Sort of 2xpMHC-multimer^+^ CD62L^+^CD45RO^+^ single cells into individual wells of a CD3/CD28 antibody coated culture plate. Performance of TCR SCAN RACE-PCR (59) on expanded single cell derived clones for TCRαβ sequence amplification. Identification of TCRαβ via Next Generation Sequencing.

**Suppl. Fig 2.**
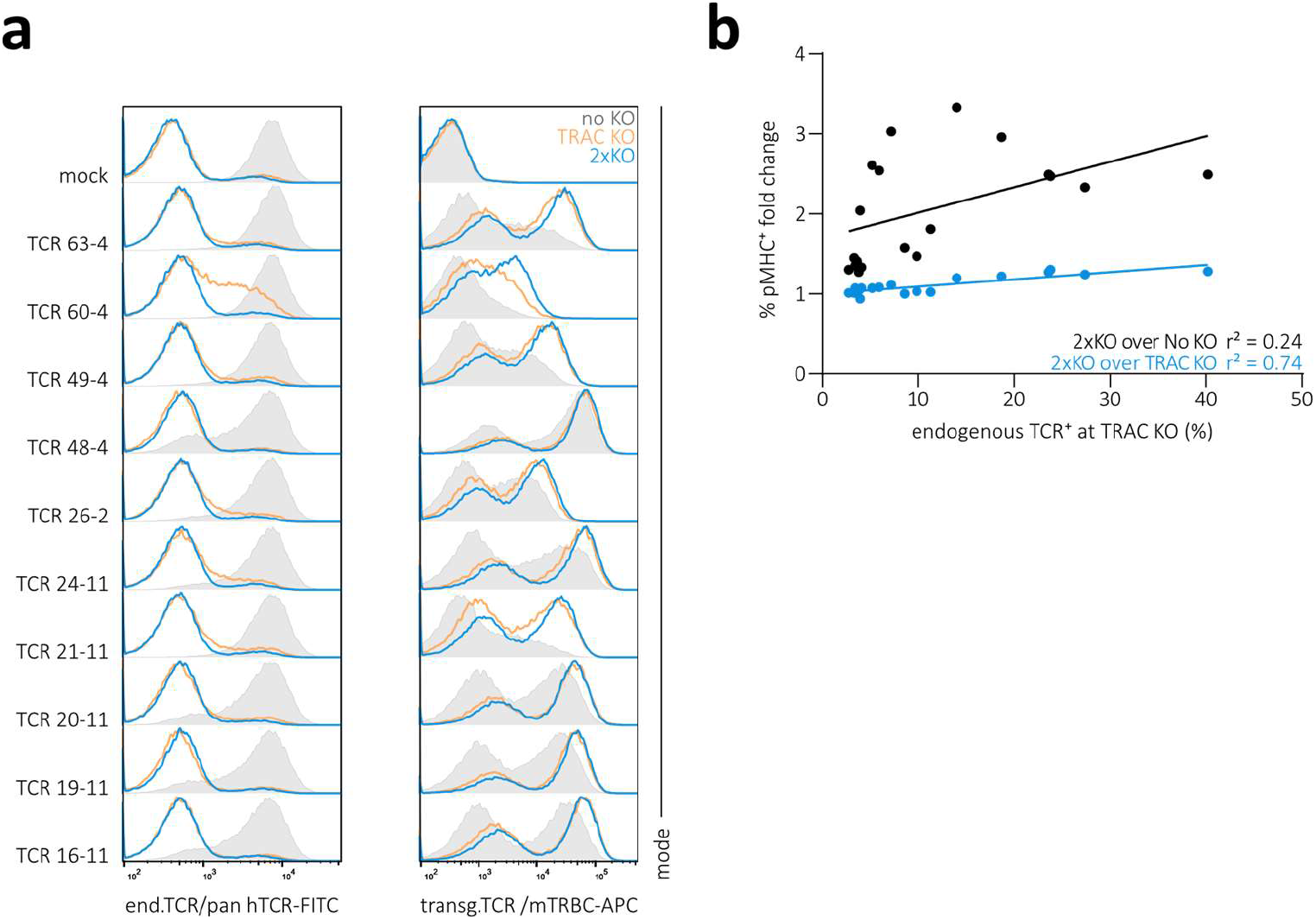
Competition and mispairing between endogenous and transgenic TCRs reduce transgenic TCR surface expression. **(a)** Expression of endogenous TCR (left) and transgenic TCR (right) of ten A1/pp50-specific retrovirally transduced TCRs in human PBMCs without additional KO of the endogenous TCR (no KO, grey), with KO of the TCR α-chain only (TRAC KO, orange) or KO of both TCR α- and β-chain (2xKO, blue). Data complement analyses shown in Fig. 1d for a total of 19 individual A1/pp50-specific TCRs. **(b)** Correlation of mispairing (as measured by percentage of endogenous TCR^+^ cells after TRAC KO) to fold change in pMHC-multimer^+^ (percentage of CD8^+^) between editing groups as indicated. Each dot represents one of 19 individual A1/pp50-specific TCRs. Fitting by linear regression.

**Suppl. Fig 3.**
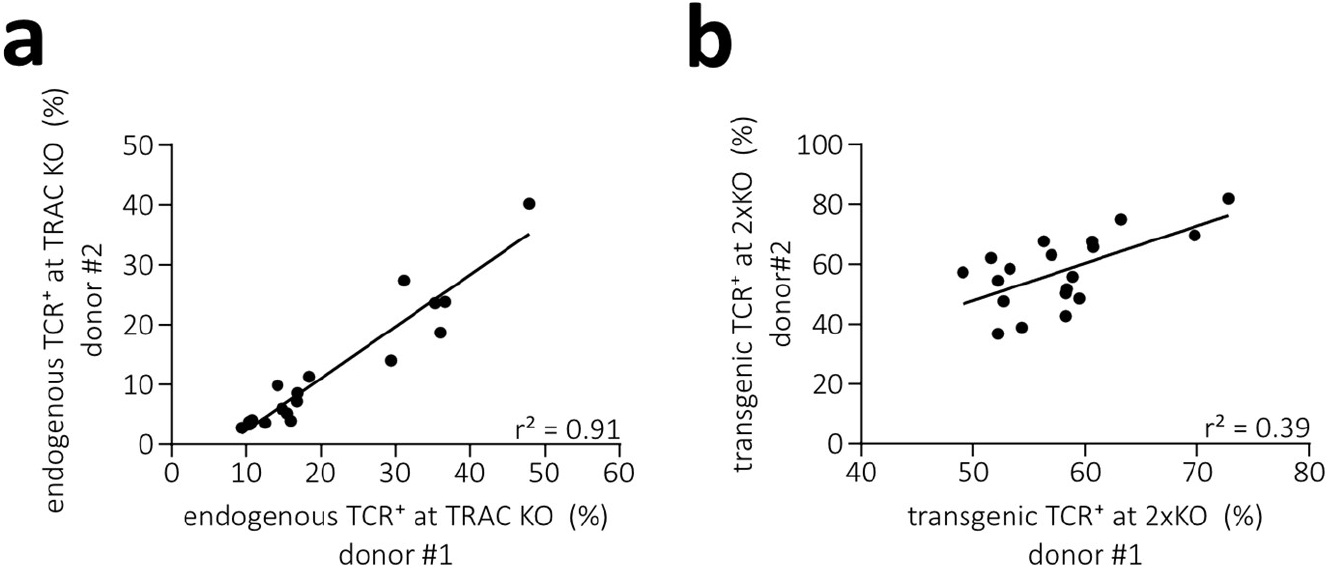
Inter-donor comparison reveals remaining variability in TCR-engineered T cell products despite removal of TCR-intrinsic mispairing through KO of the endogenous TCR. **(a)** Mispairing of transgenically expressed TCRs (as measured by percentage of endogenous TCR^+^ cells after TRAC KO) in two different donors corresponding to data shown in left panel of Fig. 1f. **(b)** Number of transgenic TCR expressing cells (percentage of CD8^+^) after 2xKO in two different donors corresponding to data shown in right panel of Fig. 1f. Each dot represents one of 19 A1/pp50-specific TCRs. Fitting by linear regression.

**Suppl. Fig 4.**
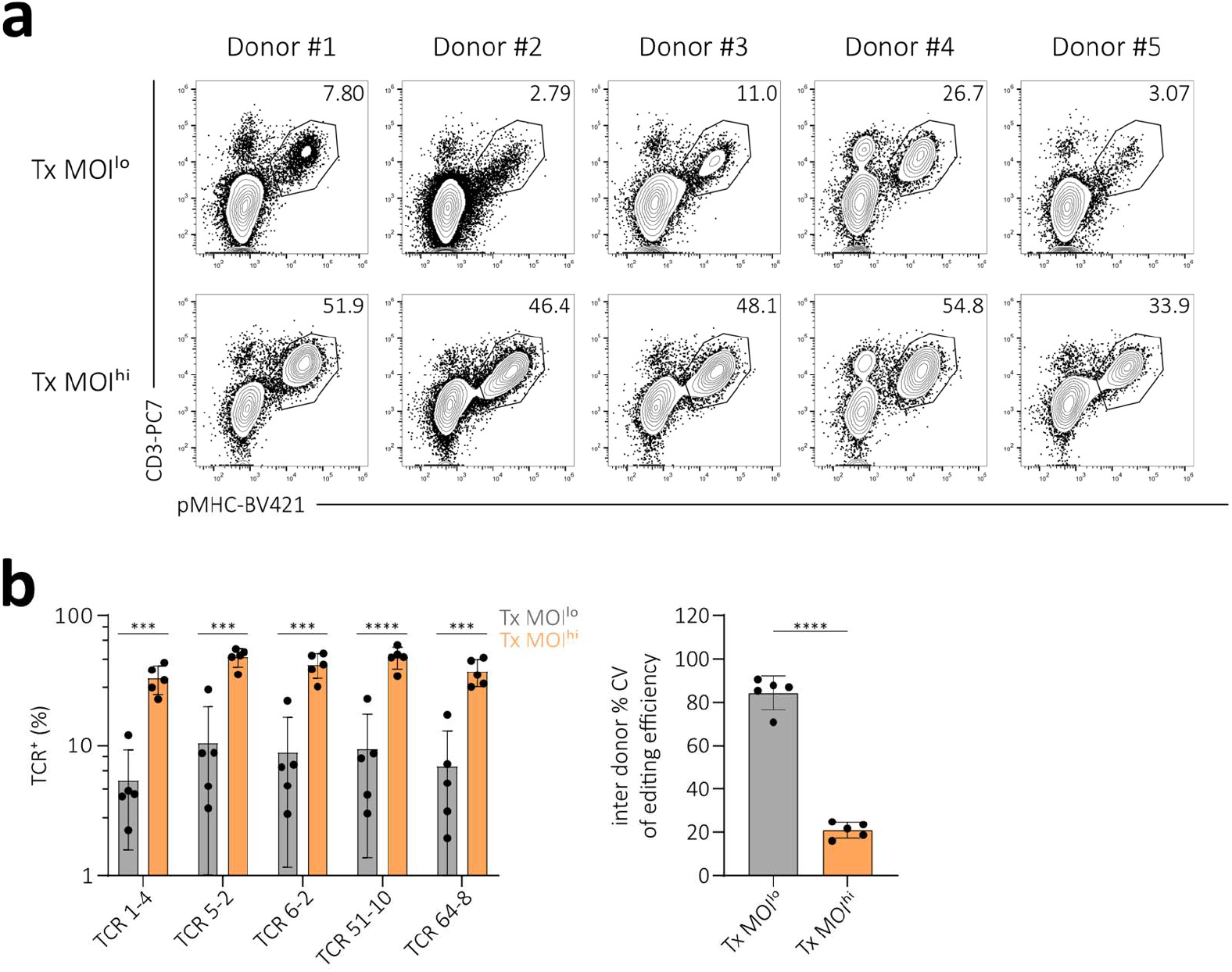
Conventional TCR editing with a low virus MOI increases inter-donor editing variability. **(a)** Editing efficiency (illustrated by pMHC-multimer/CD3 co-staining) for one representative fully human A2/pp65-specific TCR in PBMCs of five different donors. Editing was performed either via retroviral transduction with a low virus MOI or with a high virus MOI and in both cases additional elimination of the endogenous TCR. Numbers denote percentages of CD8^+^. **(b)** Quantification of efficiency of two different editing methods (retroviral transduction with a low virus MOI in grey and with a high virus MOI in orange) for five fully human A2/pp65-specific TCRs in five different donors. Each dot represents one TCR in one of five donors (left). Also shown is the quantification of inter-donor variability (as measured by coefficient of variation of editing efficiency) for the two different editing methods (right). Each dot represents inter-donor variability of one of five TCRs. Depicted is mean ± s.d.. Statistical testing by two-tailed unpaired Student’s t-test, **** p<0.0001, *** p<0.001.

**Suppl. Fig 5.**
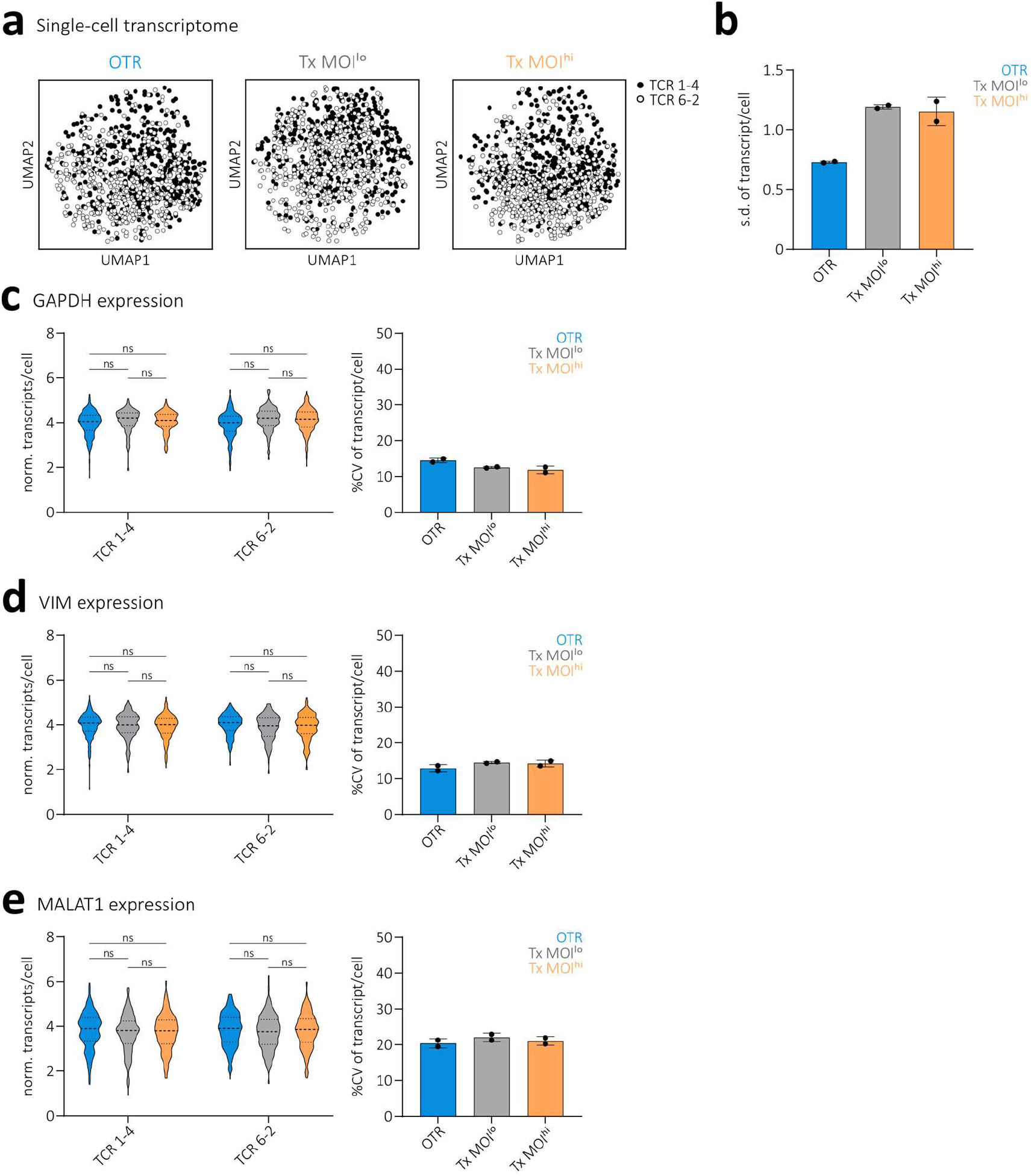
ScRNA seq data reveal differential transgenic TCR expression between OTR and conventional editing without any further transcriptomic changes. **(a)** UMAP on whole transcriptome data compared by TCR, n = 450 cells per TCR. **(b)** Standard deviation of transgenic TCR expression. Depicted is mean ± s.d., n = 256 cells per editing method. Statistical testing by one-way ANOVA (* p<0.05) followed by Tukey’s multiple comparisons test, * p<0.05. **(c**,**d**,**e)** Quantification of expression of house-keeping genes as indicated (left). Violin plots indicate frequency distribution, median and quartiles are shown with dashed lines. Statistical testing by two-way ANOVA ((c): ns for editing method, **** p<0.001 for TCR; (d): ns for editing method, * p<0.05 for TCR; (e): ns for editing method, ns for TCR) followed by Tukey’s multiple comparisons test. Quantification of cell-to-cell variability (right). Depicted is mean ± s.d., n = 256 cells per editing method.

**Suppl. Fig 6.**
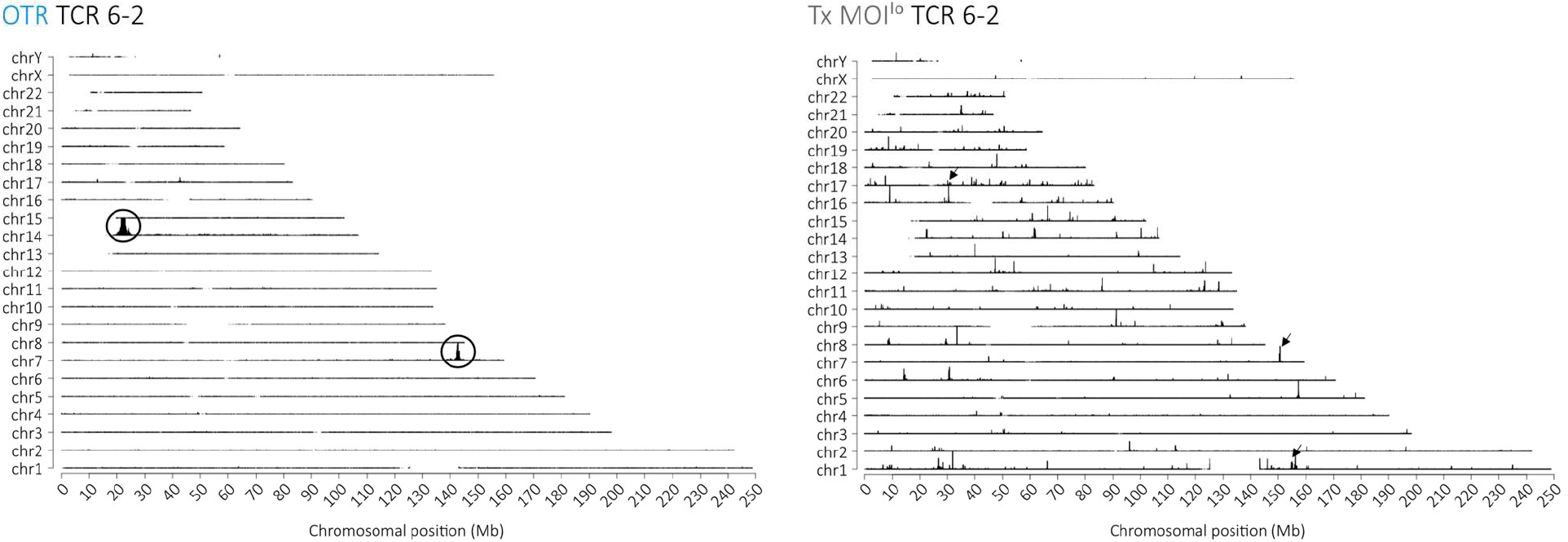
Transgene genomic integration is highly variable after conventional editing. TLA coverage across the human genome for integration of TCR 6-2. The chromosomes are indicated on the y-axis, the chromosomal position on the x-axis. Circles indicate more abundant integration sites. Arrows indicate examples of less abundant integration sites. Similar results were obtained with an independent primer set.

**Suppl. Fig 7.**
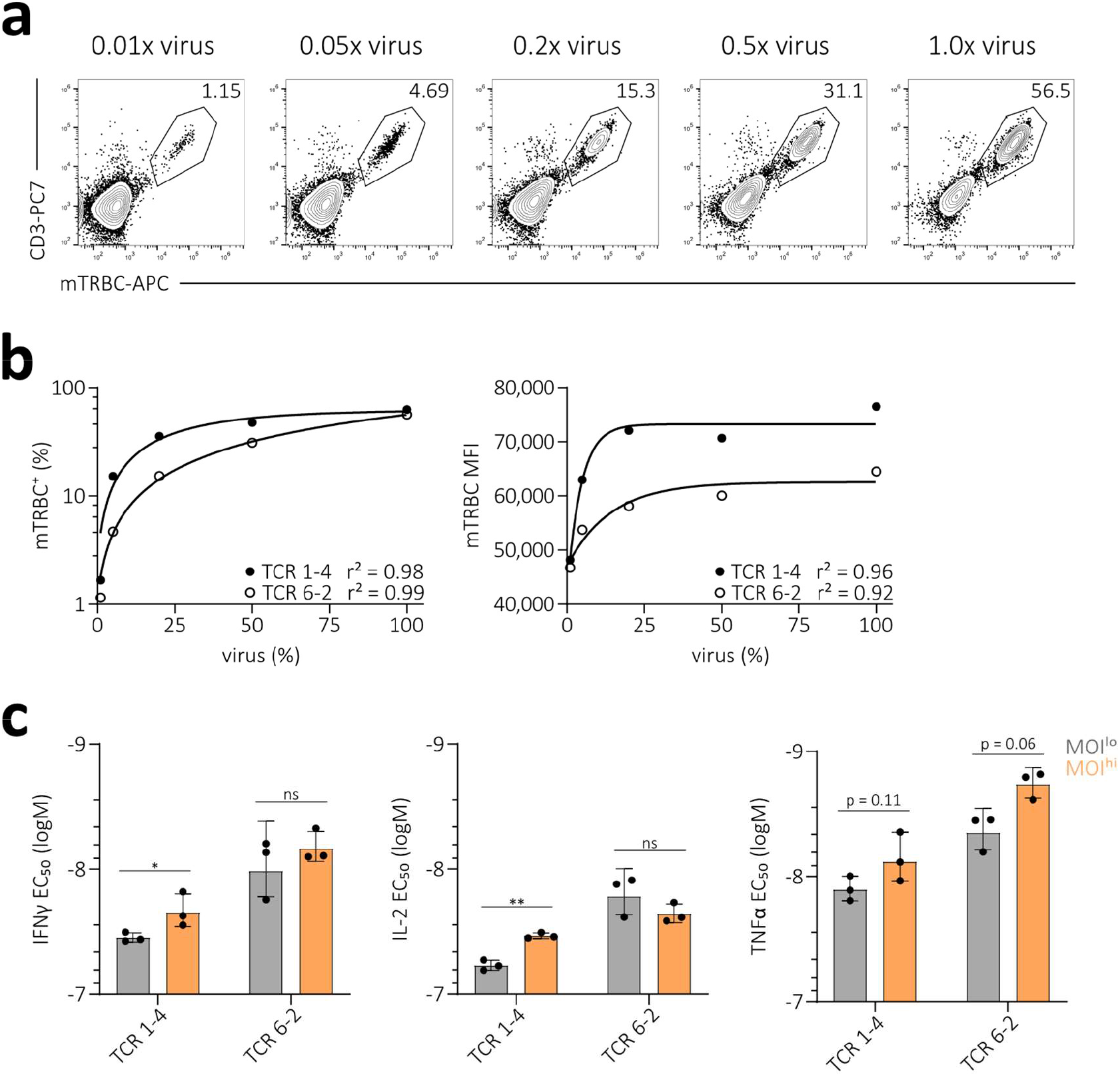
Conventional editing introduces variability in transgenic TCR surface expression and functionality. **(a)** TCR surface expression (indicated by mTRBC/CD3 co-staining) of the A2/pp65-specific TCR 1-4. Editing was performed in human PBMCs via retroviral transduction with a titration of virus MOIs (indicated as fold-dose of 1.0x virus) and additional elimination of the endogenous TCR (as indicated by CD3^-^mTRBC^-^ cells). Numbers indicate editing efficiencies as percentage of CD8^+^. **(b)** Correlation of virus dose (as percentage of 1.0x virus) to editing efficiency (percentage mTRBC^+^ of CD8^+^) (left) and to TCR surface expression (mTRBC MFI; right). Each dot represents one of two indicated TCRs. Fitting by non-linear regression. **(c)** Half-maximum effective concentration (EC_50_) quantification of IFN-γ, IL-2 and TNFα release after antigen-specific stimulation of two A2/pp65-specific TCR-transgenic flow cytometry-sorted T cell products that were generated with a low (grey) or high (orange) virus MOI (0.01x virus and 1.0x virus respectively, see (a) and Fig. 3a). Depicted are individual replicates and mean ± s.d. Statistical testing by two-tailed unpaired Student’s t-test, ** p<0.01, * p<0.05.

**Suppl. Fig 8.**
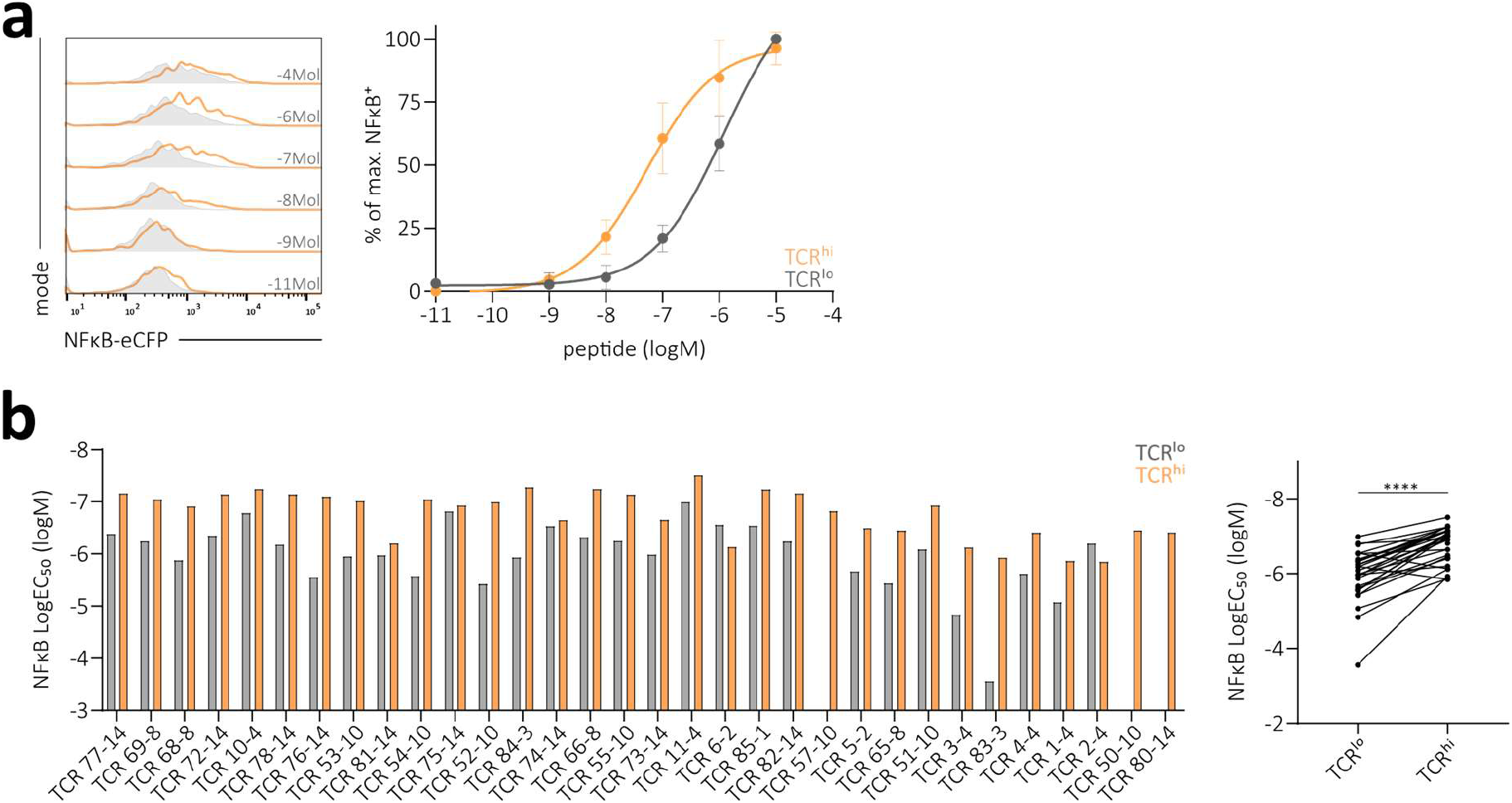
TCR surface expression levels affect peptide sensitivity of 32 A2/pp65-specific TCRs re-expressed in a TCR signaling reporter cell line. **(a)** NFκB reporter activity of cells with low (grey) and high (orange) surface expression of one representative TCR after antigen-specific stimulation with different peptide concentrations as indicated (left). Corresponding EC_50_ curves (logM, log molar; right). **(b)** Quantification of NFκB signal log molar EC_50_ values of 32 A2/pp65-specific TCRs (left). TCRs are ordered from left to right according to NFκB E_max_ shown in Fig. 3d. For each TCR, NFκB signal LogEC_50_ is shown for cells with low (grey) and high (orange) TCR surface expression. Each bar represents the mean of three replicates. Direct comparison of NFκB signal EC_50_ between cells with low and high TCR surface expression for all 32 A2/pp65-specific TCRs (right). Statistical testing by two-tailed paired Student’s t-test, **** p<0.0001.

**Suppl. Fig 9.**
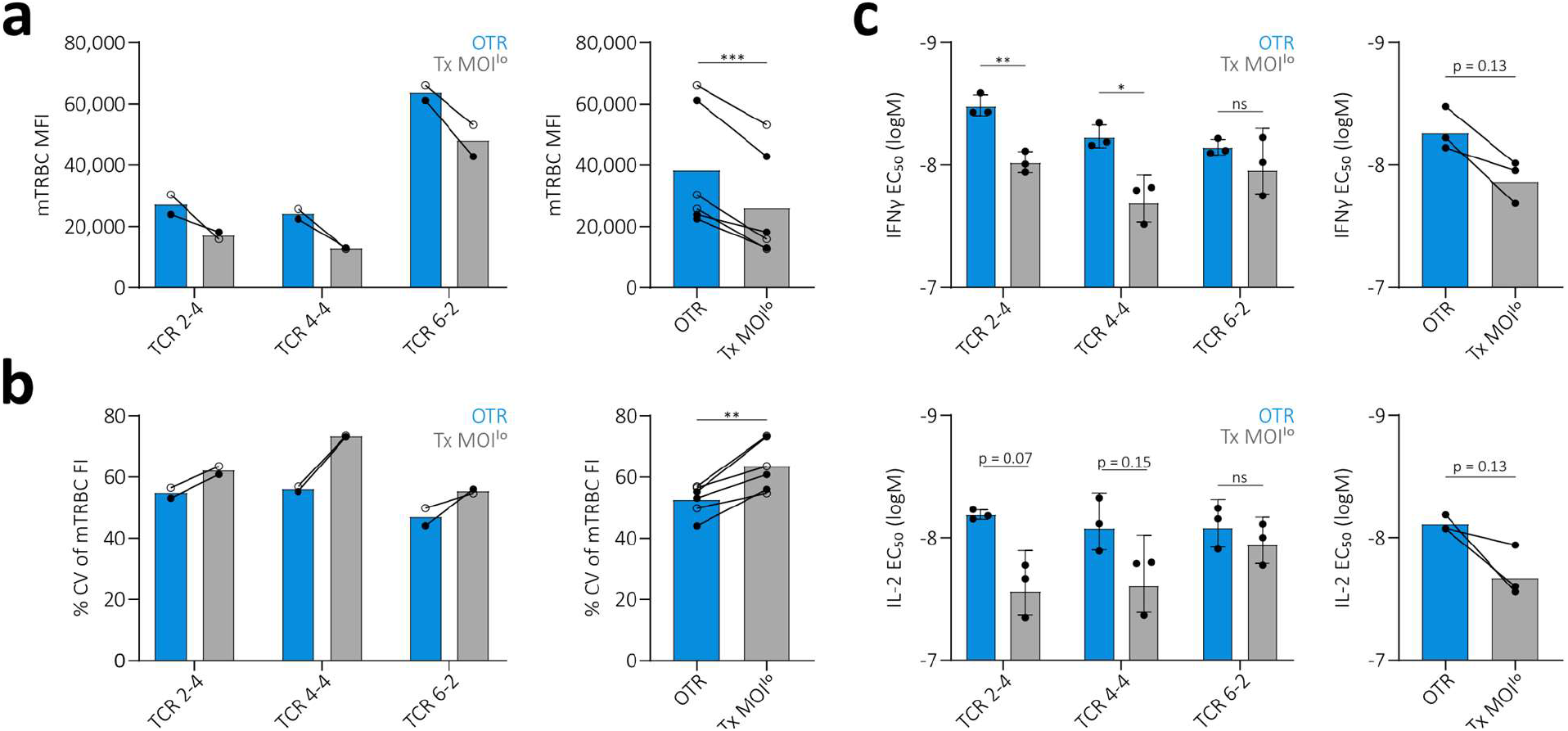
OTR generates T cell products with increased functionality compared to conventionally edited low-virus dose transduced T cells. **(a)** TCR surface expression of three A2/pp65-specific TCRs after flow cytometry sorting and two weeks of *in vitro* culture. Editing was performed in human PBMCs (derived from a different donor than depicted in Fig. 4) via OTR (blue) or retroviral transduction with a low virus MOI (grey) and in both cases additional elimination of the endogenous TCR. TCR surface expression levels (measured by mTRBC MFI) for each individual TCR at two different time points (left) and pooled for comparison of editing groups (right). **(b)** Cell-to-cell TCR surface expression variability (coefficient of variation of mTRBC FI) of T cells shown in (a) for each individual TCR at two different time points (bottom left) and pooled for comparison of editing groups (bottom right). (a,b) Each dot represents measurement of one TCR at one of two time points (day 10 after sort indicated by open circles, day 14 after sort indicated by filled circles). Statistical testing by two-tailed paired Student’s t-test, *** p<0.001, ** p<0.01. **(c)** Functional cytokine response of T cell products shown in (a). Quantification of IFNγ (top left) and IL-2 (bottom left) EC_50_ values of four individual A2/pp65-specific TCRs. Depicted are replicates and mean ± s.d.. Statistical testing by two-tailed unpaired Student’s t-test, ** p<0.01, * p<0.05. Direct comparison of IFNγ and IL-2 EC_50_ between editing groups (right). Here, each dot represents the mean of three replicates for one TCR. Statistical testing by two-tailed paired Student’s t-test.

**Suppl. Fig 10.**
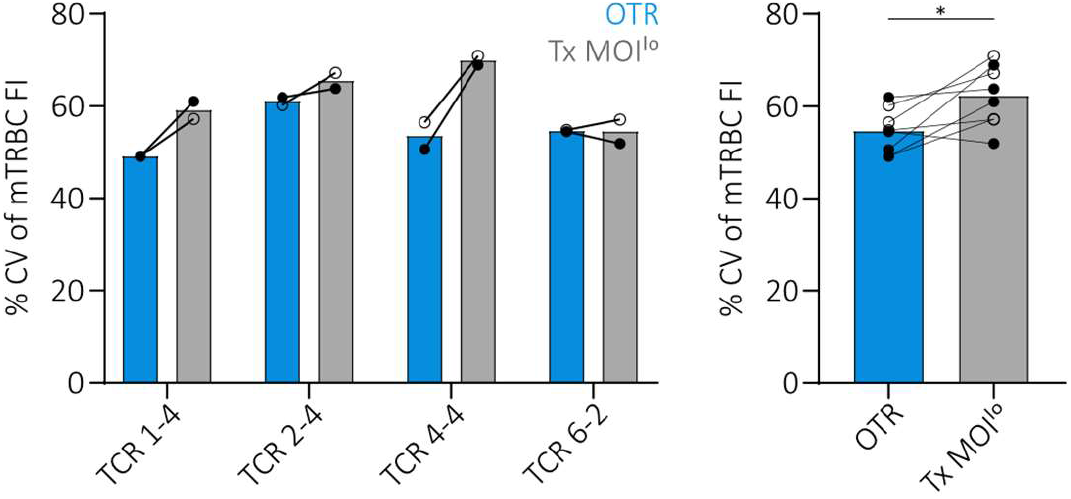
OTR with murinized TCRs generates T cell products with more homogenous TCR surface expression compared to conventional editing. Analysis of cell-to-cell TCR surface expression variability (as coefficient of variation of mTRBC FI) from TCR-transgenic T cell products shown in Fig. 4a (left). Direct comparison of TCR surface expression variability between editing methods (right). Each dot represents measurement of one TCR at one of two time points (day 10 after sort indicated by open circles, day 14 after sort indicated by filled circles). Statistical testing by two-tailed paired Student’s t-test, * p<0.05.

**Suppl. Fig 11.**
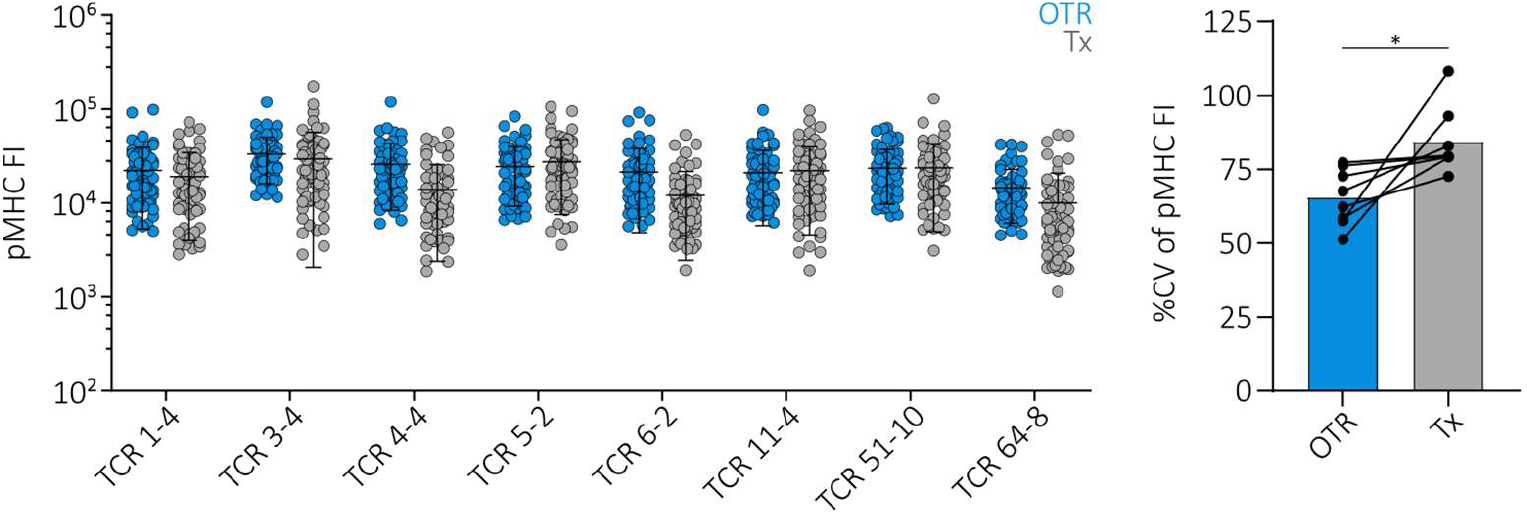
OTR with fully human TCRs generates T cell products with more homogenous TCR surface expression compared to conventional editing. pMHC-multimer FI of eight fully human A2/pp65-specific TCRs. Editing was performed either via OTR (blue) or retroviral transduction (grey) and additional elimination of the endogenous TCR (accounts for both methods). Each dot represents one single cell. For each TCR and editing method, 100 randomly drawn cells derived from a larger population are displayed to account for equal population sizes (left). Quantification of the corresponding cell-to-cell pMHC-multimer FI variability (coefficient of variation; right). Here, each dot represents the variability within a 100-cell population of one TCR. Statistical testing by two-tailed paired Student’s t-test, * p<0.05.

**Suppl. Fig 12.**
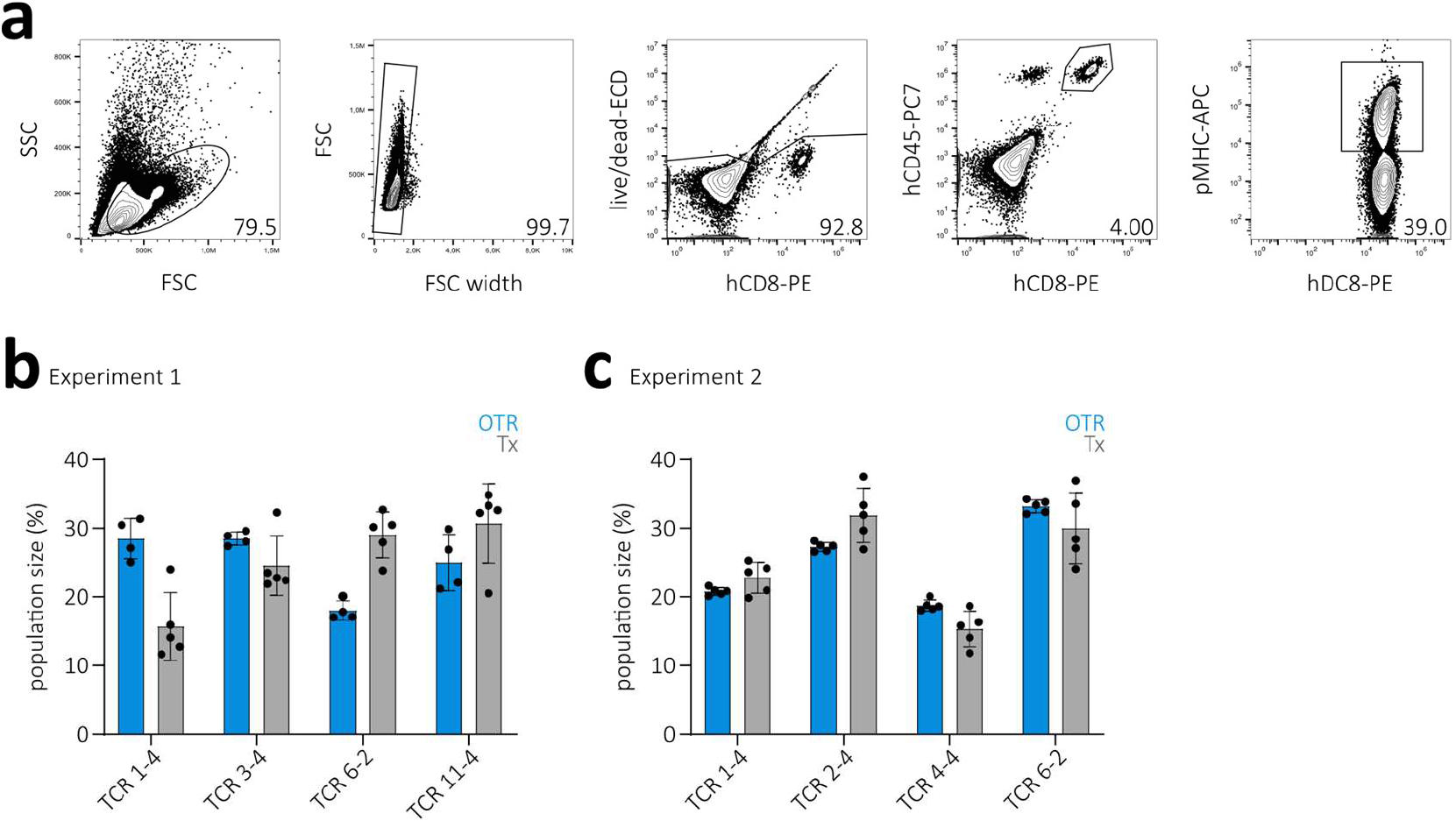
OTR generates T cell products with more reliable *in vivo* functionality. **(a)** Gating strategy for analysis of TCR-transgenic T cell responses at day 8 in liver of sacrificed mice. Numbers indicate percentage of previous gate. **(b)** Quantification of TCR-transgenic T cell responses (indicated by percentage of adoptively transferred T cells) per TCR and editing method of experiment 1 from Fig. 5e. Each dot represents one TCR in one mouse. Bars indicate mean recruitment ± s.d.; n = 4-5 mice per group. **(c)** As in (b), but for experiment 2 (also see Fig. 5f); n = 5 mice per group.

